# Routine Labs in Hospital Patients: Iatrogenic Anemia and Missed Acute Kidney Injury

**DOI:** 10.1101/2024.01.16.24301361

**Authors:** Dawson Dean

**Affiliations:** Univeristy of Kentucky

**Keywords:** Choosing Wisely, Acute Kidney Injury, Hospital Acquired Anemia

## Abstract

Guidelines recommend avoiding unnecessary laboratory tests to minimize risks of anemia in hospitalized patients as well as reduce costs. Avoiding routine labs, however, may introduce new risks of missing conditions that do not have physical exam or history findings, such as Acute Kidney Injury. This study analyzes retrospective data for routine laboratory results and simulates different strategies for skipping labs. It estimates potential benefits from avoiding iatrogenic anemia as well as costs from increased risk of Acute Kidney Injury. In a simplified estimate of pure dollar costs, the costs of daily labs appear to significantly outweigh the costs of missing Acute Kidney Injury, but there are costs to skipping routine labs.

## Introduction

The laboratory test is the microscope of the internist, and illuminates processes such as renal arteriole blood flow or distal tubule transporter activity that are invisible to the gross findings of a physical exam or history. However, the choosing Wisely guidelines recommend clinicians “Don’t perform laboratory blood testing unless clinically indicated so that the risk of iatrogenic anemia is minimized.” (Mueller MM, 2019). Further, the Society of Hospital Medicine Choosing Wisely guidelines recommend,

“Additionally, reducing the frequency of daily unnecessary phlebotomy can result in significant cost savings for hospitals” (Society of Hospital Medicine Medicine – Adult Hospital, n.d.) There are, however, risks to not monitoring a patient’s laboratory values from conditions such as Acute Kidney Injury (AKI), but the guidelines leave open the question of what defines clinically indicated and do not specify the costs and savings of routine lab tests. Like many medical decisions, choosing how frequently to monitor laboratory values benefits from comparing costs and benefits of routine lab draws, and this paper attempts to quantify these risks and benefits.

## Methods

This study is a retrospective analysis of 48,204 admissions at University of Kentucky Hospitals, which are teaching hospitals serving central and eastern Kentucky. All patients were admitted to general Internal Medicine Teams between 2015 and 2019 inclusive. All patients were at least 18 years old, had at least three days of laboratory values collected, and had an estimated Glomerular Filtration Rate (eGFR) that exceeded 20 at least once.

All patients were de-identified, and all data analysis was performed with Python software running on Python 3.7.0 using standard numerical libraries. The source code for all analysis is open source and may be downloaded from: http://www.dawsondean.com/publicSrcCode/ddeanAnemiaAndAKISrcCode.tar

### Methods – Acute Kidney Injury

Acute Kidney Injury (AKI) is defined by the KDIGO guidelines: (Khwaja, 2012)

- KDIGO I: Creatinine increase by 0.3 mg/dl within 48 hours or Increase in Creatinine to 1.5-1.9 times baseline or Urine volume <0.5 ml/kg/hr for 6-12 hours
- KDIGO 2: Creatinine increase to 2.0-2.9 times baseline or Urine Output < 0.5ml/kg/hr for at least 12hrs
- KDIGO 3: Creatinine increase to 2.0-2.9 times baseline or Urine Output < 0.3ml/kg/hr for at least 24hrs or Anuria for at least 12hrs

In all of these criteria, the baseline Creatinine is assumed to be have occurred within the prior 7 days. The current study omits the restriction of seven days, and instead considers the baseline to be the lowest Creatinine measured during the admission prior to each new lab. Additionally, because Urine output is often not recorded reliably, this study only uses changes in Creatinine to define AKI. This study also limits the criteria of Creatinine rising 0.3mg/dL to patients whose previous Creatinine was less than 1.7mg/dL to avoid false positives from normal variation in advanced CKD. This study also ignores any patient whose estimated Glomerular Filtration Rate (GFR) never rises above 20 mL/min, because such patients may be on hemodialysis. All GFR values are computed using the CKD-Epi Equation (Levey, 2010)

### Methods - AKI and Skipping Labs

The question of whether skipping lab values will lead to missed AKIs is difficult because there is no way to describe the results of laboratory tests that were never performed. An approximation is to use “Virtual Admissions”, which are any sequence of 3 or more hospital days with daily labs. There may be several virtual admissions in one actual admission, and each sequence of daily labs is considered a separate virtual admission (Figure 1).

**Figure 1.**
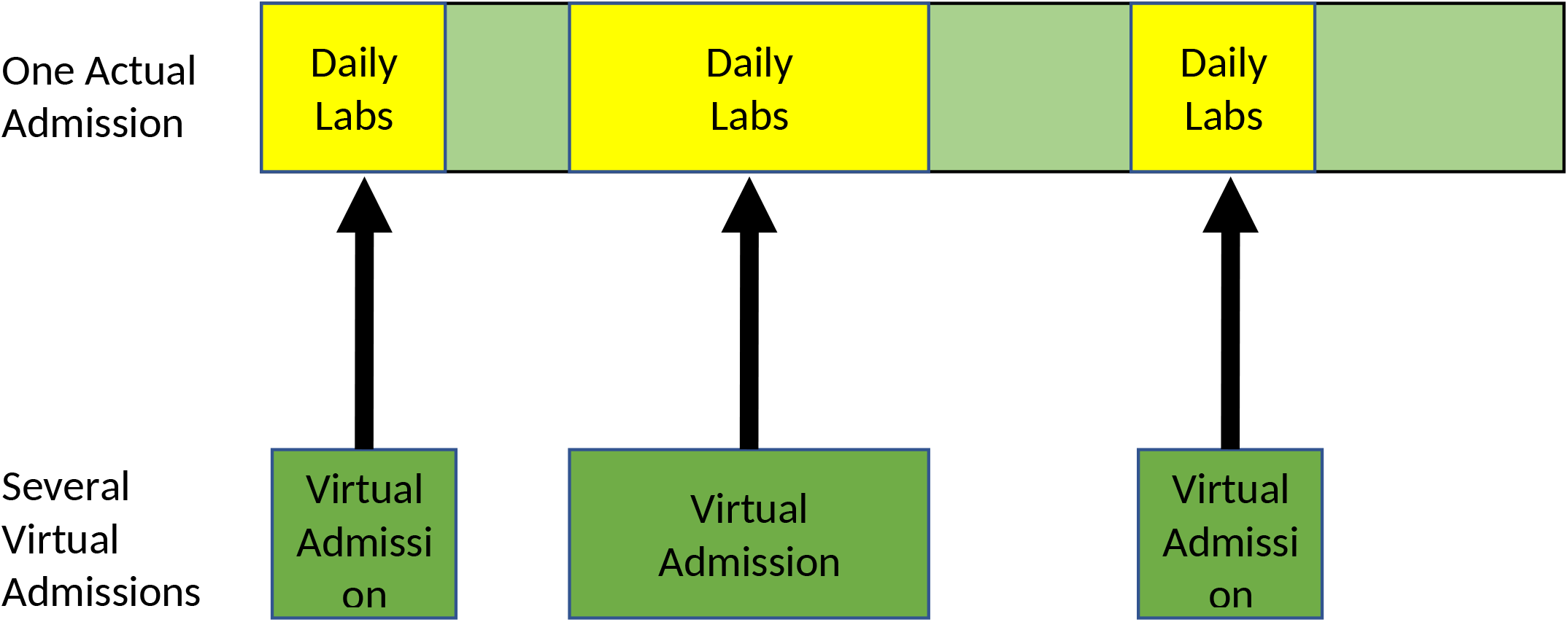

This study simulates skipping labs by ignoring some values and estimates sensitivity by comparing the number of AKIs observed with and without skipping labs:

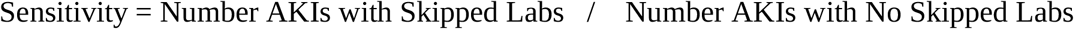

Ideally, we skip enough labs to avoid unnecessary blood draws, but not so many labs that we miss episodes of AKI. There are many methods to make this tradeoff and several were tested in this study, including:

- Randomly skipping labs starting on the first day of a hospitalization
- Randomly skipping labs after the fourteenth day of a hospitalization (emphasizes stable patients who remain inpatient due to placement issues)
- Randomly skipping labs only when Creatinine has been stable - the most recent Creatinine is between 0.7 and 1.3x of the 3-day running average.

### Methods – Anemia

The WHO definition of anemia is absolute Hemoglobin below 13.0 g/dL for Men and 12.0 g/dL for Women (World Health Organization, 2011), but this difference can also be calculated in several ways. Some papers investigating anemia (Salisbury, 2010) (Koch, 2013) use the difference between Hemoglobin on admission and discharge, and only consider patients with normal Hemoglobin on admission. This approach, however, excludes acute on chronic anemias that happen during the middle of an admission. Instead, this study expands the WHO definitions to use comparable drops in Hemoglobin between the highest and lowest values recorded during the hospital admission:

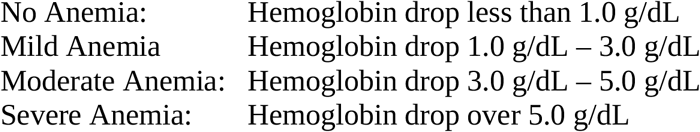

Additionally, anemia can also happen after a transfusion, so this study defines Peak Hemoglobin as:

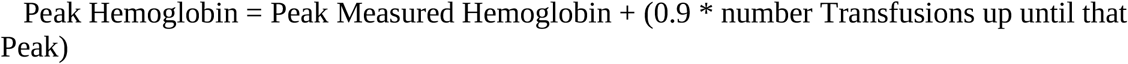

This Hemoglobin drop will be designated as Peak-To-Trough, while the drop in Hemoglobin measured only at admission and discharge will be designated as Admission-to-Discharge. When the type of anemia is not specified, it will be Peak-To-Trough.

## Results

This study is a retrospective analysis of 48,204 admissions at University of Kentucky Hospitals who were admitted between 2015 and 2019 inclusive.

### Results – Acute Kidney Injury

For AKI analysis, this study examined 43,683 patient admissions at the University of Kentucky between 2015 and 2019 with an eGFR that was at least 20 at some time during the admission. Among these non-dialysis hospital admissions, 18.2% (n=7,935) had any AKI, 12.7% (n=5,566) had a KDIGO-1 AKI, 3.6% (n=1,568) had a KDIGO-2 AKI, and 1.3% (n=801) had a KDIGO-3 AKI. Significant Elixhauser statistics of University of Kentucky patients with AKI in the Hospital are described in Supplemental Table 2. Diagnoses like Hypertension and Valvular Cardiomyopathy seem to be more associated with higher levels of kidney Injury. However, only 46% of patients with KDIGO 3 AKI based on their Creatinine had an Elixhauser ICD code related to AKI.

Most cases of KDIGO 2 and KDIGO 3 AKI were present on admission (79% of KDIGO 2 and 75% of KDIGO 3), while only 52% of KDIGO 1 AKI happened before admission so 48% of KDIGO 1 AKI happened after admission (Supplemental Figure 16). Most patients who developed AKI in the hospital had a baseline Creatinine between 0.5 and 0.75, and many patients with AKI were receiving nephrotoxic medication - 15% were prescribed a Non-Steroidal Anti-Inflammatory Drug (NSAID), 46% received Vancomycin, and 41% were on a diuretic (Supplemental Figure 18).

The test for Creatinine, however, has limits in reproducibility and possibly accuracy. Occasionally, patients in the hospital have identical labs drawn at nearly the same time, and two labs drawn within fifteen minutes apart should be very similar. Comparing the results from these near simultaneous tests suggests the reproducibility of the overall test, and may be indicative of its accuracy. The University of Kentucky uses the Roche Cobas 8000/702 (Jaffee method) assay for creatinine, and 254 pairs of Creatinine labs were collected within fifteen minutes apart. Of these near simultaneous lab pairs, only 43% of results were identical, but 90% of pairs were within a range of -0.3 and 0.3 apart (Figure 5).

### Results - AKI and Skipping Labs

We estimate the sensitivity for detecting AKI with and without daily labs by using virtual admissions. There were 11,116 Non-ESRD Virtual Admissions, and the average Virtual Length of Stay was 8.25 days. A comparable number of AKIs were found in Actual and Virtual Admissions (Supplemental Figure 26).

Random Skipping labs will miss AKIs in a dose-dependent relationship with the probability of skipping labs (Figure 4). Similarly, randomly skipping labs only after 14 days in the hospital will also miss AKIs, but misses fewer of the more severe AKIs. Supplemental Figure 25 shows the sensitivity as a function of the probability of skipping labs. Finally, skipping labs only when recent Creatinine values have been stable will also miss AKIs. However, fewer labs are skipped, and so fewer AKIs are missed. Supplemental Figure 24 shows the sensitivity as a function of the probability of skipping labs.

### Results - Hemoglobin

Of the initial patient population, 29,009 admissions had three or more Hemoglobins collected and were used for analysis. The average Hemoglobin drop (Peak-To-Trough) was 1.64 g/dL, while average Hemoglobin drop (Admission-to-Discharge) was 0.35 g/dL. Classifying anemia by Peak-To-Trough drop detected 9,762 more cases of mild anemia, 6,166 more cases of moderate anemia, and 3,654 more cases of severe anemia than Admission-to-Discharge (Supplemental Figure 1). All other statistics will use Peak-To-Trough Hemoglobin drops.

The most common Elixhauser statistics (Project, Healthcare Cost and Utilization, n.d.) (Project, Healthcare Cost and Utilization, n.d.) for patients based on ICD codes are described in Supplemental Table 1. As expected, diagnoses of Deficiency Anemia and Coagulopathy are more common in patients with anemia based on lab values, but only approximately 20% of patients with anemia based on lab values also had ICD codes for anemia.

Many patients already had some chronic anemia on admission: 11% had hemoglobin below 8, 9% below 9, 13% below 11, 13% below 12 and 12% below 13 (Supplemental Figures 2 and 3). However, anemia occurs both on admission and during the hospitalization. For example, 11% of admissions had severe anemia on admission, but 19% of admissions had severe anemia at some point during the hospitalization (Supplemental Figures 3 and 4).

Anemia that develops in the hospital tends to be slow and progressive, and patients typically don’t have large changes in hemoglobin in the middle of an admission. The average daily changes in Hemoglobin are negative and range between -0.07 g/dL to -0.2 g/dL daily after day 2 in the hospital (Supplemental Figure 12). This drop in Hemoglobin does not frequently rise to the level requiring transfusions. For example, the number of transfusions declines with both length of stay and with the number of labs collected, (Supplemental Figures 13 and 14).

However, Hemoglobin testing in daily hospital practice may not be accurate enough to draw precise conclusions. The University of Kentucky uses the Hemoglobin Sysmex XN9000 assay for measuring serum Hemoglobin, and in the data collected there were 770 pairs of tests drawn within fifteen minutes apart. Only 17% of near simultaneous lab pairs had perfect agreement, and the error range to include 90% of the collected labs was -0.8 g/dL to 0.8 g/dL, while the error range to include 95% of tests was between -1.5 g/dL to 1.5 g/dL. (Figure 2).

**Figure 2.**
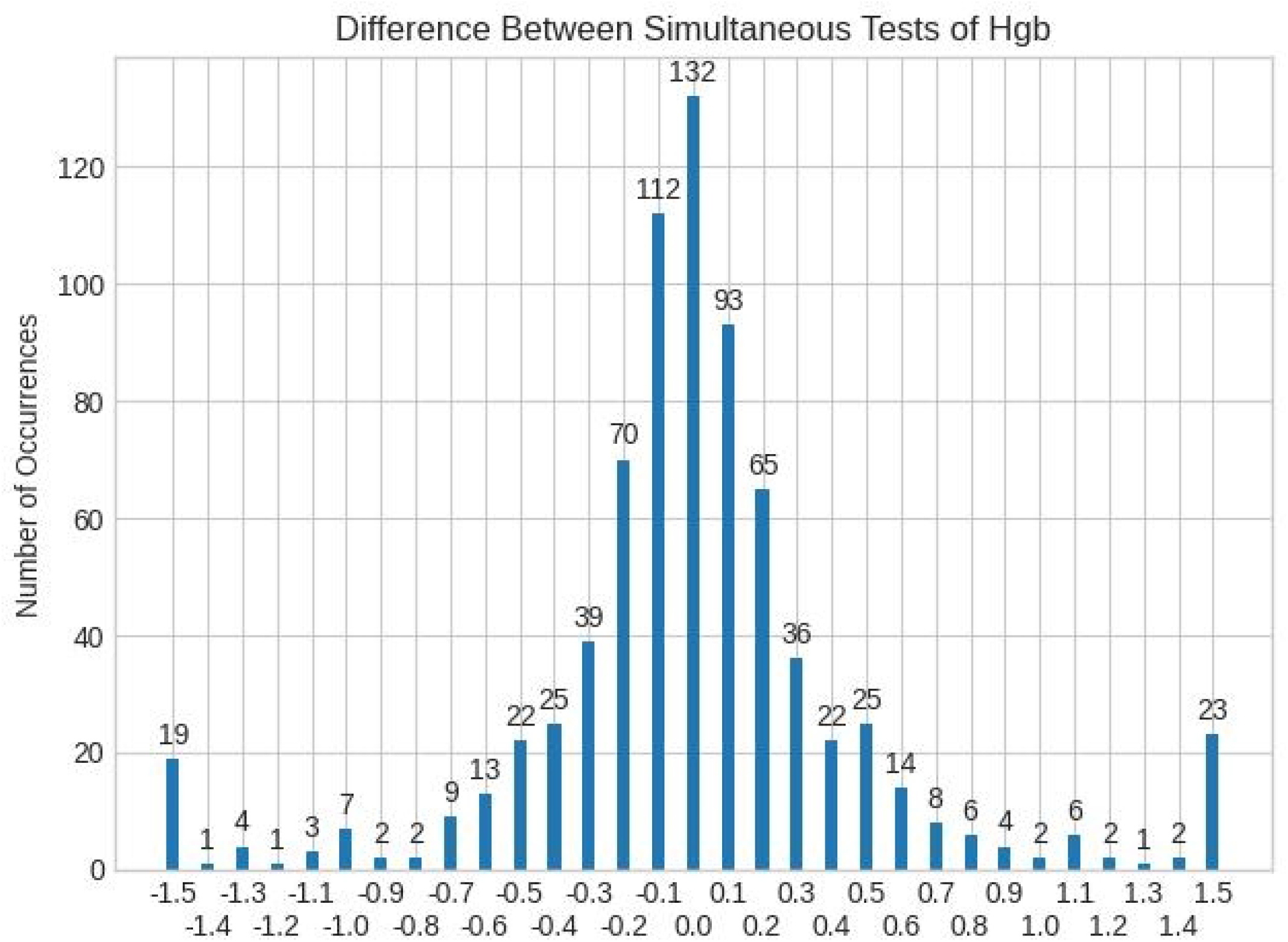

**Figure 3.**
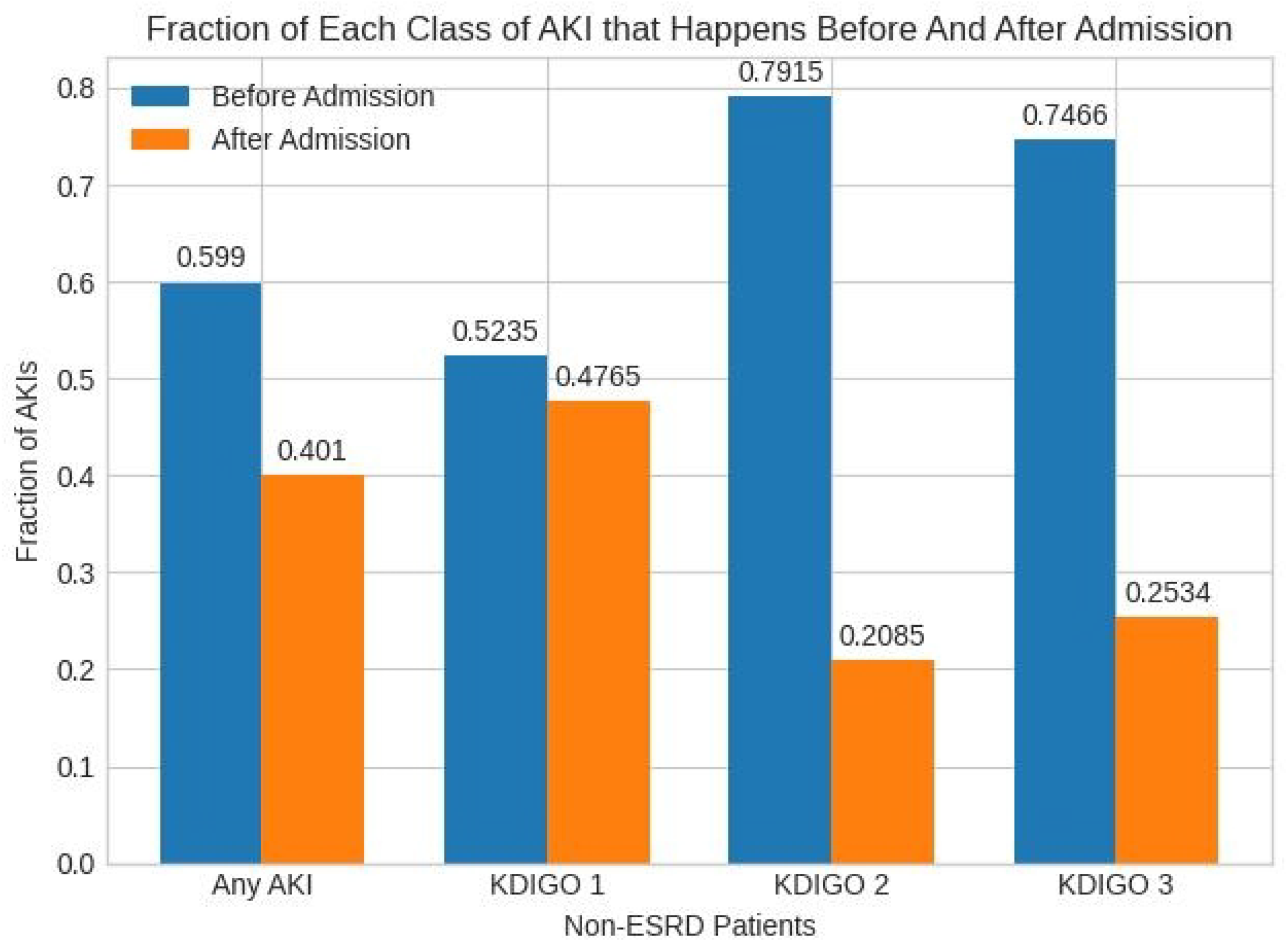

**Figure 4.**
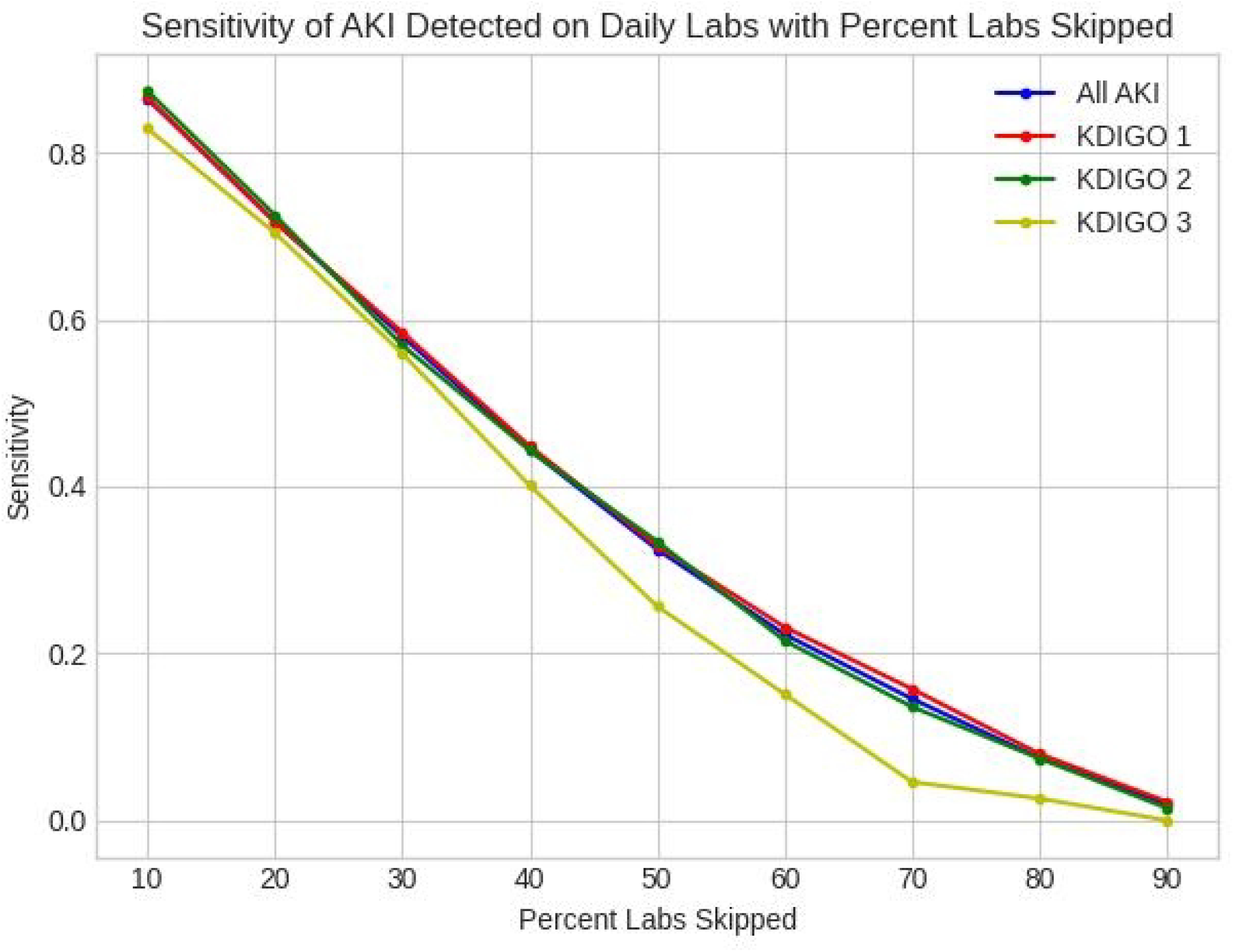

**Figure 5.**
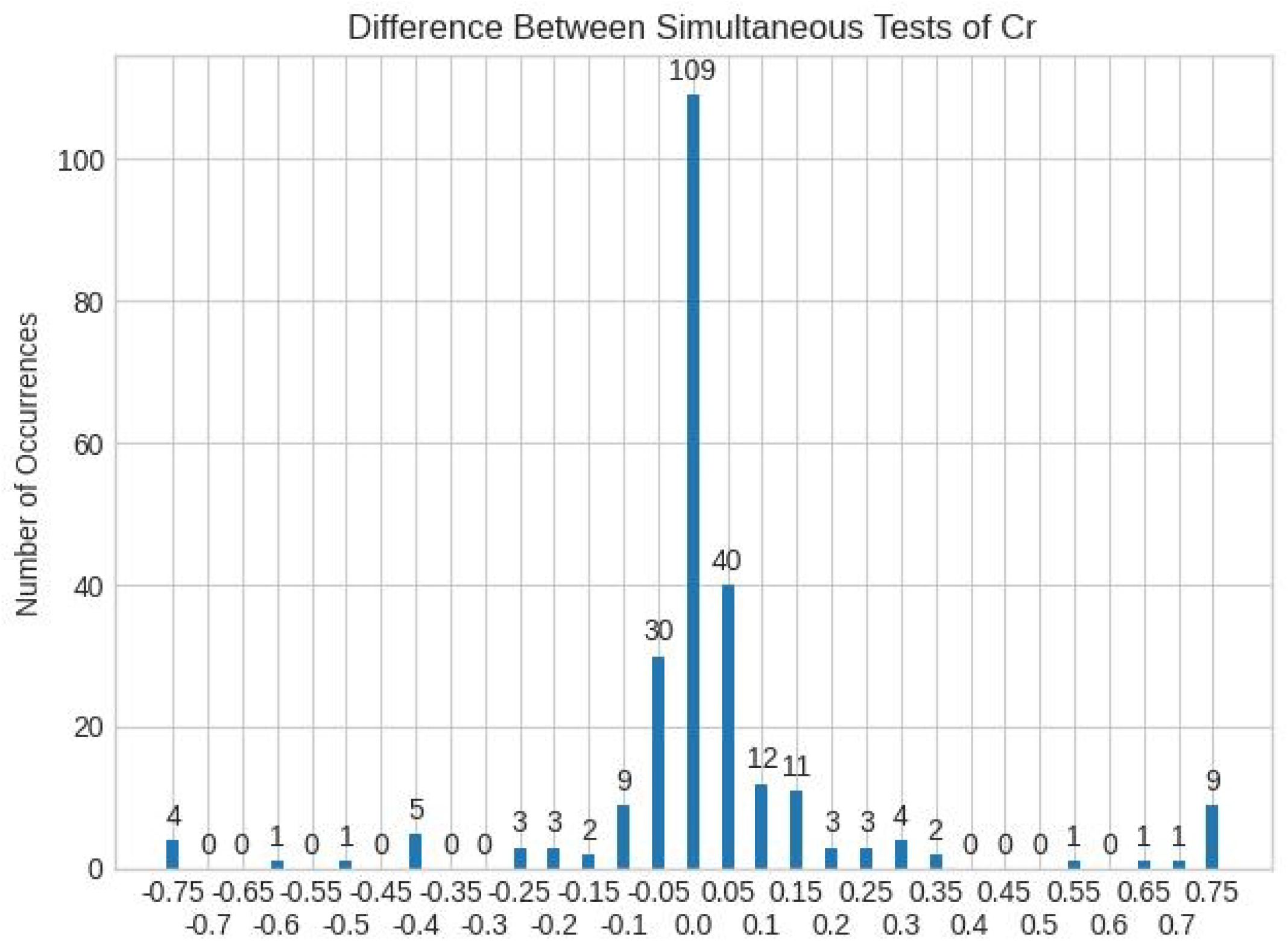

The cause of anemia while in the hospital is likely multifactorial. Lab draws is likely one cause, and the Spearman Correlation between Hemoglobin Decline and the total number of Complete Blood Count (CBC) or Basic Metabolic Panel (BMP) labs collected is 0.8116, while the correlation between Hemoglobin drop and length of stay is 0.6335. Additionally, the Spearman Correlation of Hemoglobin Decline with Platelet Decline is 0.7922, which suggests a loss of whole blood and not a limited underproduction of red blood cells. As with Hemoglobin, the Spearman Correlation between Platelets Decline and the number of labs collected is 0.8084 and between Platelets Decline and the Length of Stay is 0.6789.

The Spearman coefficient between the number of surgeries and drop in Hemoglobin is 0.4352, and the Spearman coefficient between number of Endoscopies or Colonoscopies and drop in Hemoglobin is 0.2013. Nonetheless, 62% of patients with severe anemia and 40% of patients with moderate anemia also underwent surgery (Supplemental Figures 10 and 11). Similarly, the Spearman coefficient between the highest White Blood Cell count and Hemoglobin Decline is 0.5033, while the Spearman coefficient between Hemoglobin decline and receiving antibiotics including Vancomycin is 0.3608. (Supplemental Figures 8 and 9).

## Discussion

We model the effects of reducing lab utilization by 25%, and the interested reader may use an online calculator to try different policies and assumptions http://www.dawsondean.com/CostOfRoutineLabs.html

### Costs of Skipping Labs

We will consider only the effects of skipping labs on Iatrogenic AKI, since AKI present on admission would not be affected by lab policy. If we skip 25% of labs then we skip the same number of Creatinine levels annually. Using a random sampling algorithm, we will miss 185 instances of KDIGO 1 AKI annually, 22 instances of KDIGO 2 AKI annually, and 14 instances of KDIGO 3 AKI annually, or a total of 222 cases of missed AKI annually.

Chertow (Chertow, 2005) estimated the costs of AKI; an increase in Creatinine over 0.5 mg/dl was associated with 6.5-fold increase in the odds of death, a 3.5-day increase in length of stay, and $8,902 in excess hospital costs. Ishani (Ishani, 2009) showed that patients with AKI and Chronic Kidney Disease (CKD) have increased risk for ESRD relative to patients without kidney disease (hazard ratio 41.2), and patients with AKI but no previous CKD also have increased risk for ESRD (hazard ratio 13.0).

Admission to the University of Kentucky Hospital costs $1,687 per day. If each AKI adds 3.5 additional days to the admission, then there is a total cost of $1,315,498. The cost is $1,127,569 if each AKI adds 3 additional days to the admission, $751,713 if each AKI adds 2 days, and $375,856 if each AKI adds 1 day.

### Cost-Benefit Comparison

The benefits of skipping labs is the cost savings from skipped labs as well as the avoided impacts of anemia due to those labs:

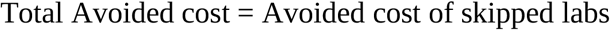

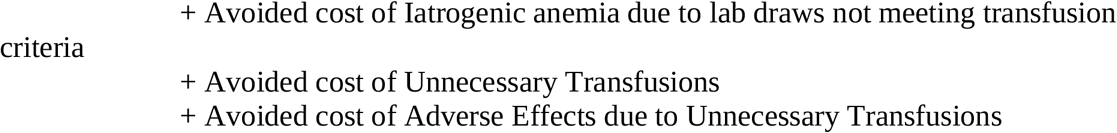

We assume each CBC costs $184, each BMP costs $187, and each transfusion costs $1,900. Reducing lab draws by 25% would provide a saving that is the sum of $5,133,470 in saved labs + $1,795,500 for avoided transfusion costs + 711 patients have reduced 12-month mortality

In this simplified estimate of pure dollar costs, we save significantly more from reduced lab costs than we would gain by detecting AKI earlier, at least partly because iatrogenic AKI seems to be relatively uncommon. However, this calculation makes several assumptions such as reducing routine labs by 25% will also reduce transfusions and anemias by the exact same percentage. The real lesson is that there are both benefits and costs to skipping labs. Quality improvement efforts often focus only on the costs in dollars and milliliters of blood of skipped labs, but what we measure affects how we behave, and we should just as carefully track the costs of skipping routine labs.

Other studies have examined routine labs and iatrogenic anemia. Salisbury et al (Salisbury, 2010) studied 2909 adult patients hospitalized with myocardial infarction and found patients with moderate or severe anemia (based on discharge Hemoglobin) had higher risk of 12-month mortality. The study did not find a causal effect between lab tests and iatrogenic anemia, and noted, “Finally, these observational data do not allow us to draw conclusions about causal relationships between HAA and mortality, and it remains unclear whether HAA is a marker for or a mediator of, poor outcomes”.

Koch et al (Koch, 2013) studied 188,447 hospitalizations, and also found patients with moderate or severe anemia had higher 12-month mortality. Again, this did not draw a clear causal connection between blood draws and anemia, and the anemia may be explained by other factors.

Makam et al (Makam, 2017) studied 29,045 hospitalizations, and found higher 30-day mortality in patients with hospital anemia, but again it is not clear whether anemia was the cause of mortality or a marker for other injury. The authors stated “Thus, blood loss due to phlebotomy, 1 of the more modifiable etiologies of HAA, was unlikely to have been the primary driver for most patients who developed severe HAA.”

This paper has several strengths: it uses Virtual Admissions to estimate skipping labs while still knowing the results of those labs, and attempts to quantify both the benefits and risks of skipping labs. There are however several weaknesses. AKI is identified only by changes in Creatinine, not urine output. Any patient with a GFR that never exceeds 20mL/min is excluded because of possible hemodialysis. Simulating skipping labs was done only on patients who had daily labs, so a clinician was possibly already concerned about renal injury. This study also assumes that any Hemoglobin is collected as part of a CBC and any Creatinine is collected as part of a BMP. This study also uses the same categories of costs of anemia as other studies, but with slightly broader definitions (includes acute on chronic anemias).

For the purpose of comparing costs and benefits, there were average 52,930 CBCs collected annually, and average 57,726 BMPs collected annually.

## Supporting information

Supplemental Material - Iatrogenic Anemia and Missed Acute Kidney Injury

## Data Availability

All data produced in the present study are available upon reasonable request to the authors. Release of data will require authorization by University of Kentucky, College of Medicine, Institutional Review Board

## Acknowledgements

The project described was supported by the NIH National Center for Advancing Translational Sciences through grant number UL1TR001998. The content is solely the responsibility of the authors and does not necessarily represent the official views of the NIH.

