## Supplemental Material - Iatrogenic Anemia and Missed Acute Kidney Injury for "Routine Labs in Hospital Patients: Iatrogenic Anemia and Missed Acute Kidney Injury"

### Supplemental Figures and Tables

*Table 1 – Selected Elixhauser Properties of Patients Grouped by Anemia*

| | All Patients | No Anemia | Mild Anemia | Moderate Anemia | Severe Anemia |
|---|---|---|---|---|---|
| Deficiency Anemia | 7.0% (3580) | 5.0% (931) | 5.0% (819) | 9.0% (738) | 20.0% (1092) |
| Blood Loss Anemia | 0.0% (146) | 0.0% (28) | 0.0% (18) | 0.0% (27) | 1.0% (73) |
| Coagulopathy | 3.0% (1293) | 2.0% (332) | 2.0% (240) | 3.0% (220) | 9.0% (501) |
| Diabetes (complicated) | 1.0% (385) | 1.0% (156) | 1.0% (81) | 1.0% (78) | 1.0% (70) |
| Diabeles (uncomplicated) | 2.0% (735) | 1.0% (231) | 1.0% (202) | 2.0% (172) | 2.0% (130) |
| Heart Failure | 2.0% (747) | 1.0% (168) | 1.0% (217) | 2.0% (154) | 4.0% (208) |
| Hypertension | 29.0% (14192) | 30.0% (5711) | 25.0% (3956) | 29.0% (2293) | 41.0% (2232) |
| Liver Disease (mild) | 6.0% (2759) | 4.0% (697) | 4.0% (670) | 8.0% (622) | 14.0% (770) |
| Neuro Disorders | 9.0% (4404) | 8.0% (1424) | 7.0% (1094) | 12.0% (936) | 17.0% (950) |
| Renal failure (moderate) | 10.0% (4959) | 8.0% (1595) | 8.0% (1234) | 10.0% (817) | 24.0% (1313) |
| Valvular Cardiomyopathy | 10.0% (4919) | 7.0% (1393) | 8.0% (1206) | 13.0% (1031) | 23.0% (1289) |

| | | | | | |
|---|---|---|---|---|---|
| Weight Loss | 11.0% (5175) | 8.0% (1443) | 9.0% (1458) | 13.0% (1032) | 23.0% (1242) |

Table 2 – Selected Elixhauser Properties of Patients Grouped by AKI

| | No AKI | KDIGO 1 | KDIGO 2 | KDIGO |
|---|---|---|---|---|
| Deficiency Anemia | 6.0% (5602) | 10.0% (291) | 15.0% (192) | 16.0% ([illegible] |
| Coagulopathy | 3.0% (2258) | 3.0% (74) | 6.0% (80) | 9.0% ([illegible] |
| Diabeles (uncomplicated) | 2.0% (1350) | 3.0% (79) | 5.0% (68) | 2.0% ([illegible] |
| Heart Failure | 1.0% (1210) | 2.0% (64) | 5.0% (60) | 5.0% ([illegible] |
| Hypertension (uncomplicated) | 26.0% (22418) | 33.0% (965) | 41.0% (509) | 44.0% ([illegible] |
| Liver Disease (mild) | 6.0% (5138) | 7.0% (210) | 12.0% (146) | 10.0% ([illegible] |
| Neuro Disorders | 9.0% (7962) | 16.0% (460) | 20.0% (243) | 30.0% ([illegible] |
| Obese | 5.0% (4262) | 4.0% (122) | 8.0% (98) | 10.0% ([illegible] |
| Psychosis | 4.0% (3476) | 6.0% (161) | 5.0% (66) | 13.0% ([illegible] |
| Renal failure (moderate) | 6.0% (5084) | 12.0% (355) | 19.0% (230) | 46.0% ([illegible] |
| Valvular Cardiomyopathy | 10.0% (9012) | 16.0% (475) | 17.0% (205) | 20.0% ([illegible] |
| Weight Loss | 11.0% (9632) | 15.0% (444) | 17.0% (208) | 30.0% ([illegible] |

Figure 1: Anemia by Peak to Trough vs First to Last

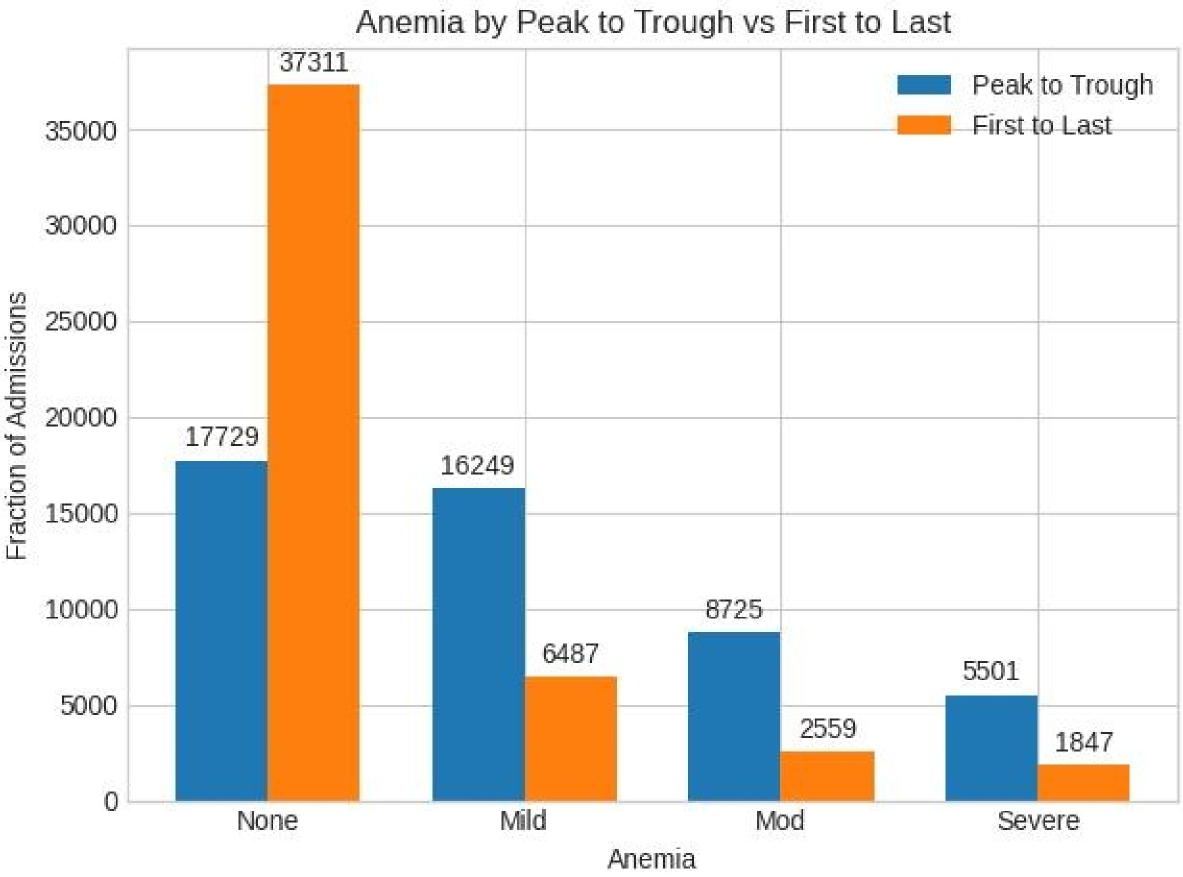

Figure 2

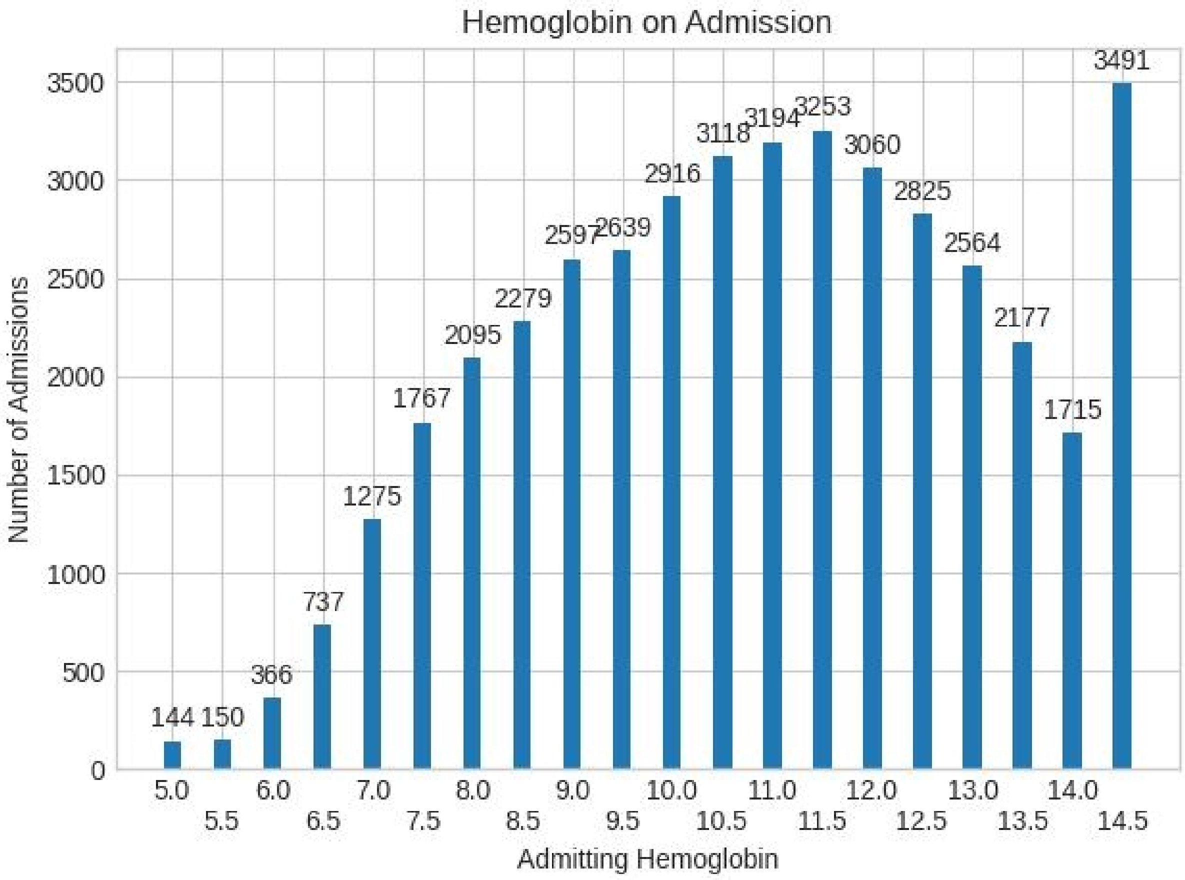
Hemoglobin on Admission
Number of Admissions
Admitting Hemoglobin
0
500
1000
1500
2000
2500
3000
3500
5.0
5.5
6.0
6.5
7.0
7.5
8.0
8.5
9.0
9.5
10.0
10.5
11.0
11.5
12.0
12.5
13.0
13.5
14.0
14.5
144
150
366
737
1275
1767
2095
2279
2597
2639
2916
3118
3194
3253
3060
2825
2564
2177
1715
3491

Figure 3

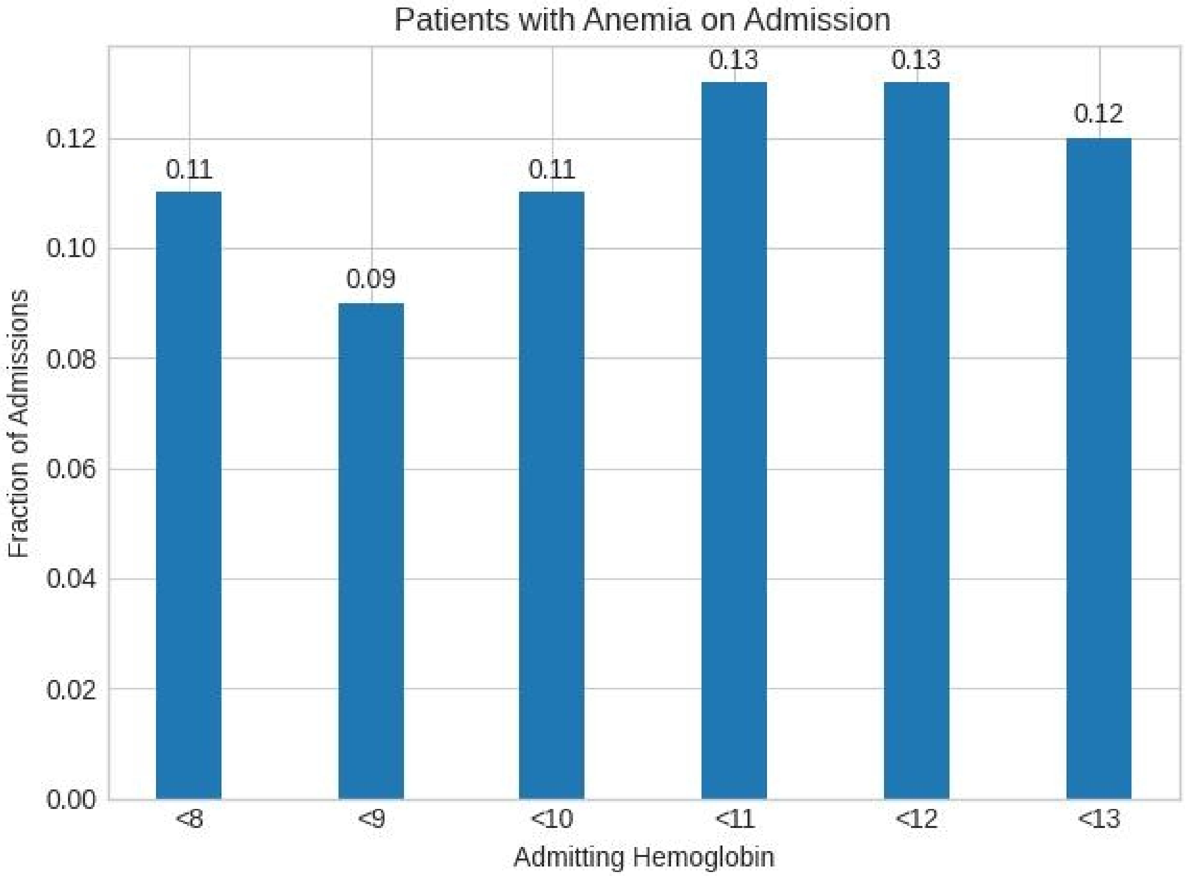

Patients with Anemia on Admission
0.11
0.09
0.11
0.13
0.13
0.12
0.12
0.10
0.08
0.06
0.04
0.02
0.00
Fraction of Admissions
<8
<9
<10
<11
<12
<13
Admitting Hemoglobin

Figure 4

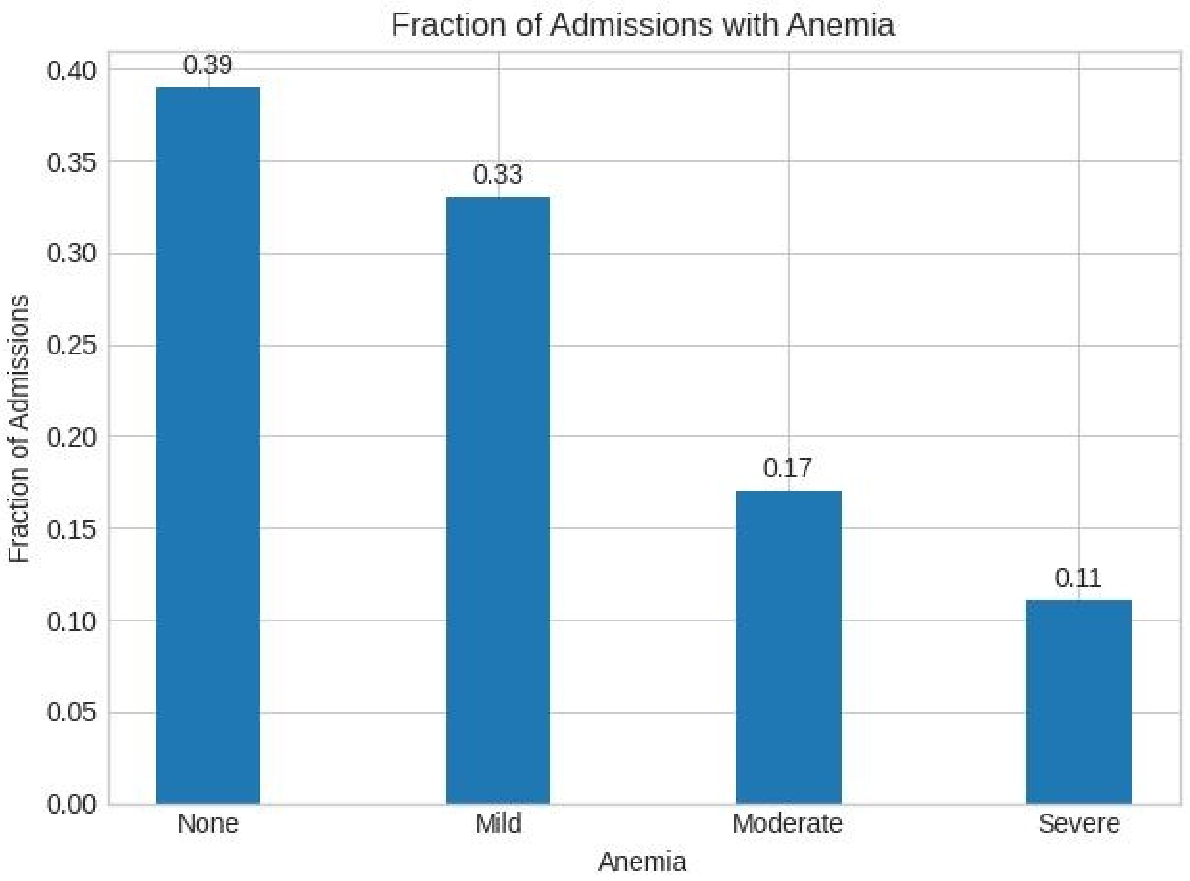
Fraction of Admissions with Anemia
Fraction of Admissions
0.00
0.05
0.10
0.15
0.20
0.25
0.30
0.35
0.40
0.39
0.33
0.17
0.11
None
Mild
Moderate
Severe
Anemia

Figure 5

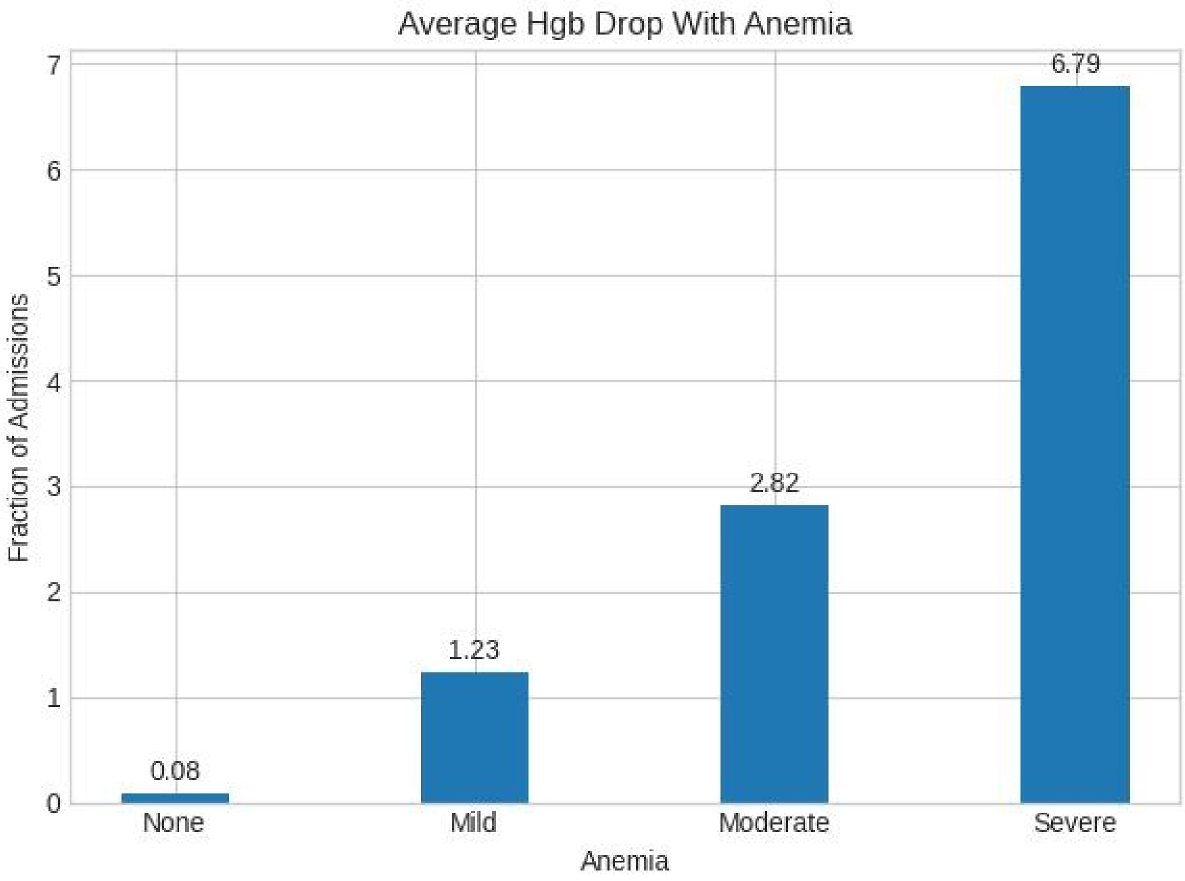
Average Hgb Drop With Anemia
Fraction of Admissions
Anemia
None
Mild
Moderate
Severe
0.08
1.23
2.82
6.79
0
1
2
3
4
5
6
7

Figure 6

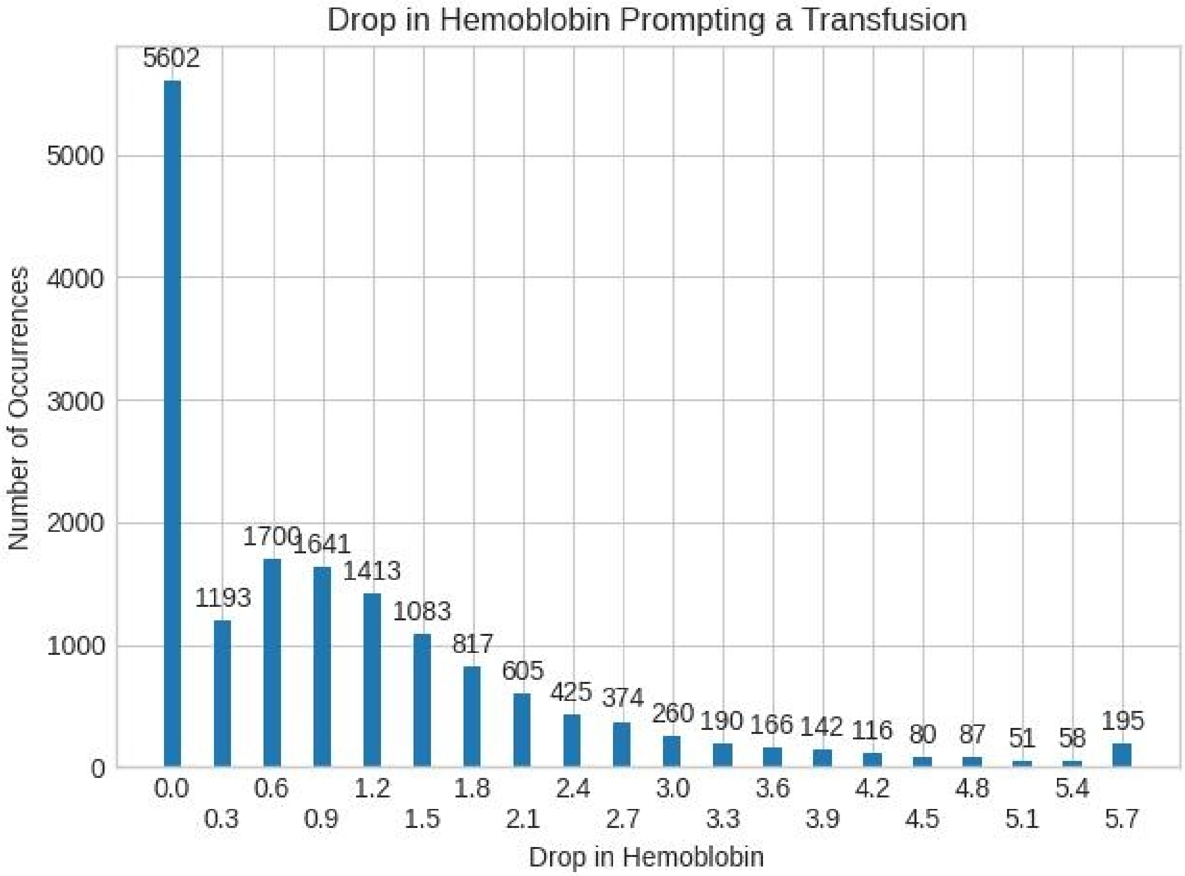
Drop in Hemoblobin Prompting a Transfusion
Number of Occurrences
0
1000
2000
3000
4000
5000
5602
1193
1700
1641
1413
1083
817
605
425
374
260
190
166
142
116
80
87
51
58
195
0.0
0.3
0.6
0.9
1.2
1.5
1.8
2.1
2.4
2.7
3.0
3.3
3.6
3.9
4.2
4.5
4.8
5.1
5.4
5.7
Drop in Hemoblobin

Figure 7

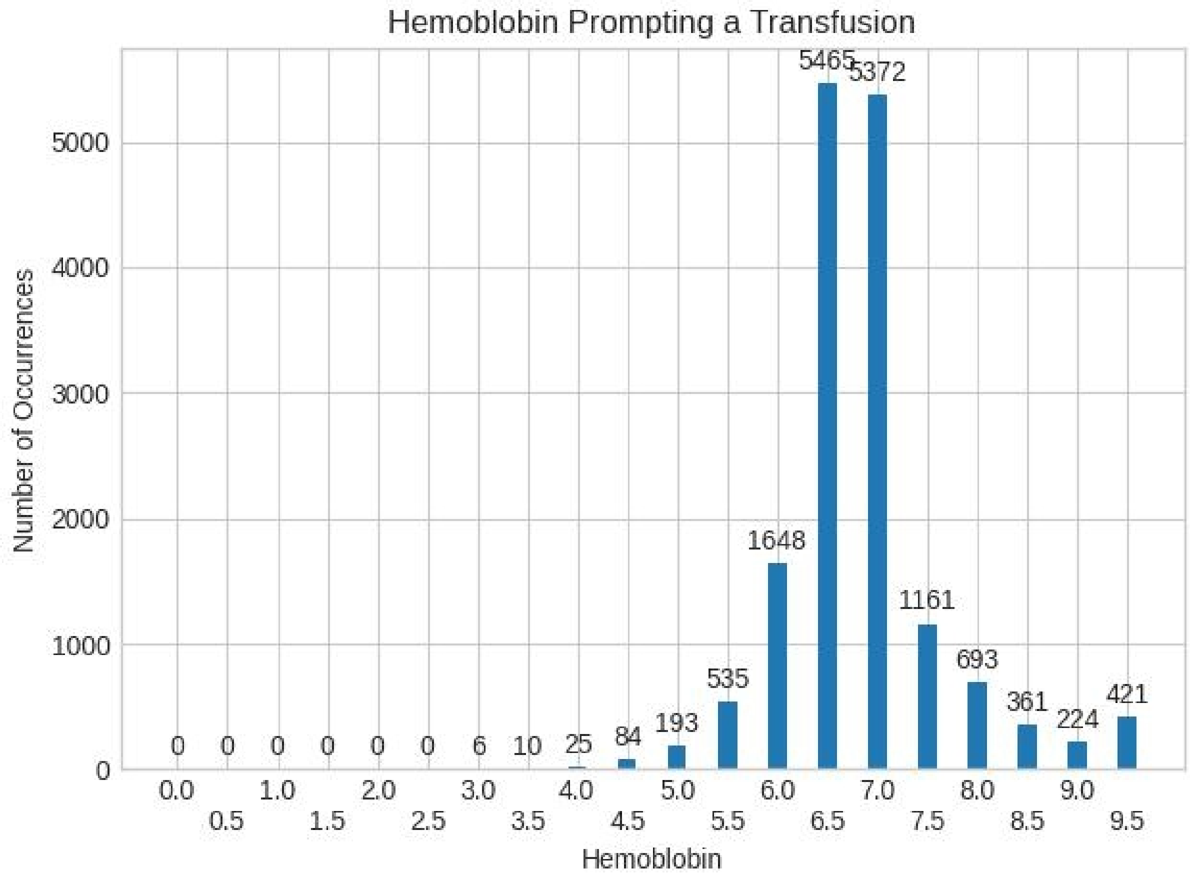

Hemoblobin Prompting a Transfusion
Number of Occurrences
Hemoblobin
0
1000
2000
3000
4000
5000
0 0 0 0 0 0 6 10 25 84 193 535 1648 5465 5372 1161 693 361 224 421
0.0 0.5 1.0 1.5 2.0 2.5 3.0 3.5 4.0 4.5 5.0 5.5 6.0 6.5 7.0 7.5 8.0 8.5 9.0 9.5

Figure 8 Fraction of Admissions with Anemia and IV Antibiotics

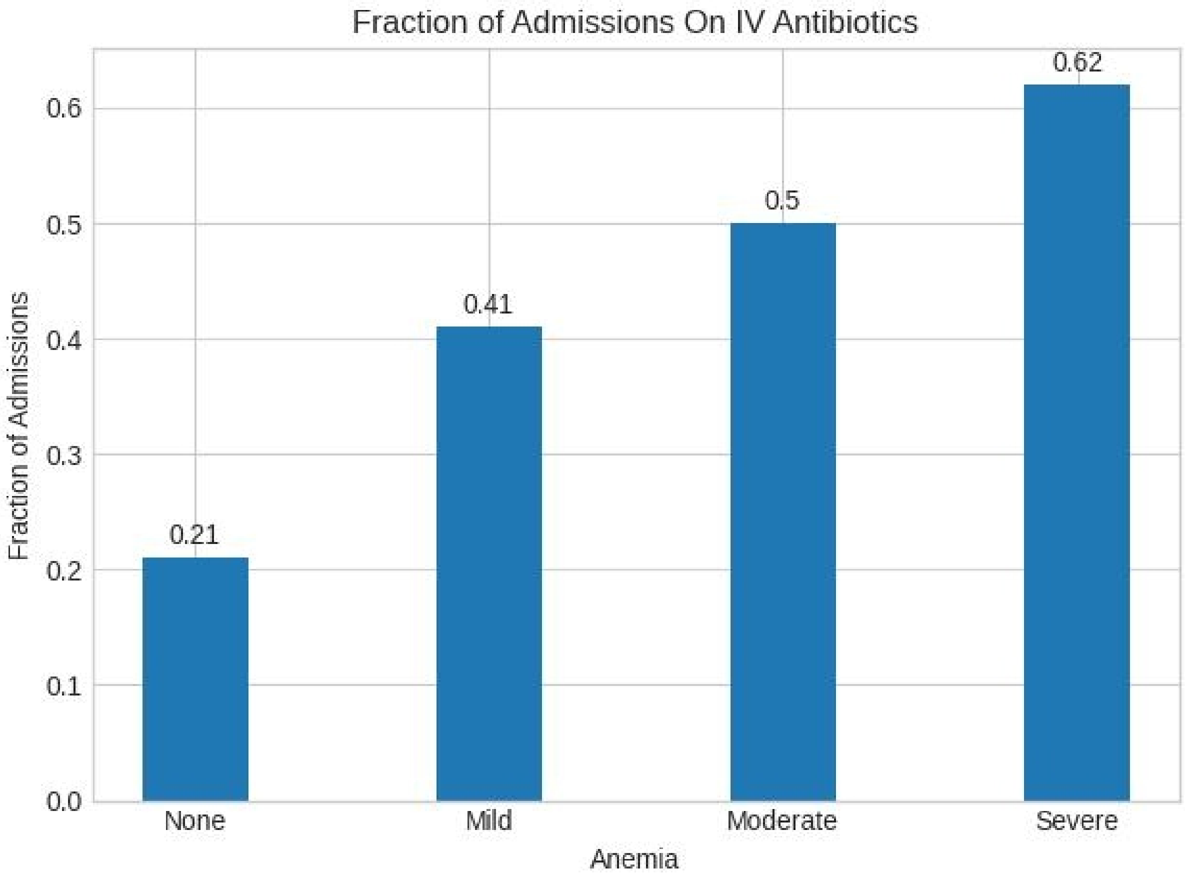


This is fraction of patients with each level of Hemoglobin Drop (not fraction of all admissions)

Figure 9 – Fraction Admissions with Anemia and Peak WBC over 12

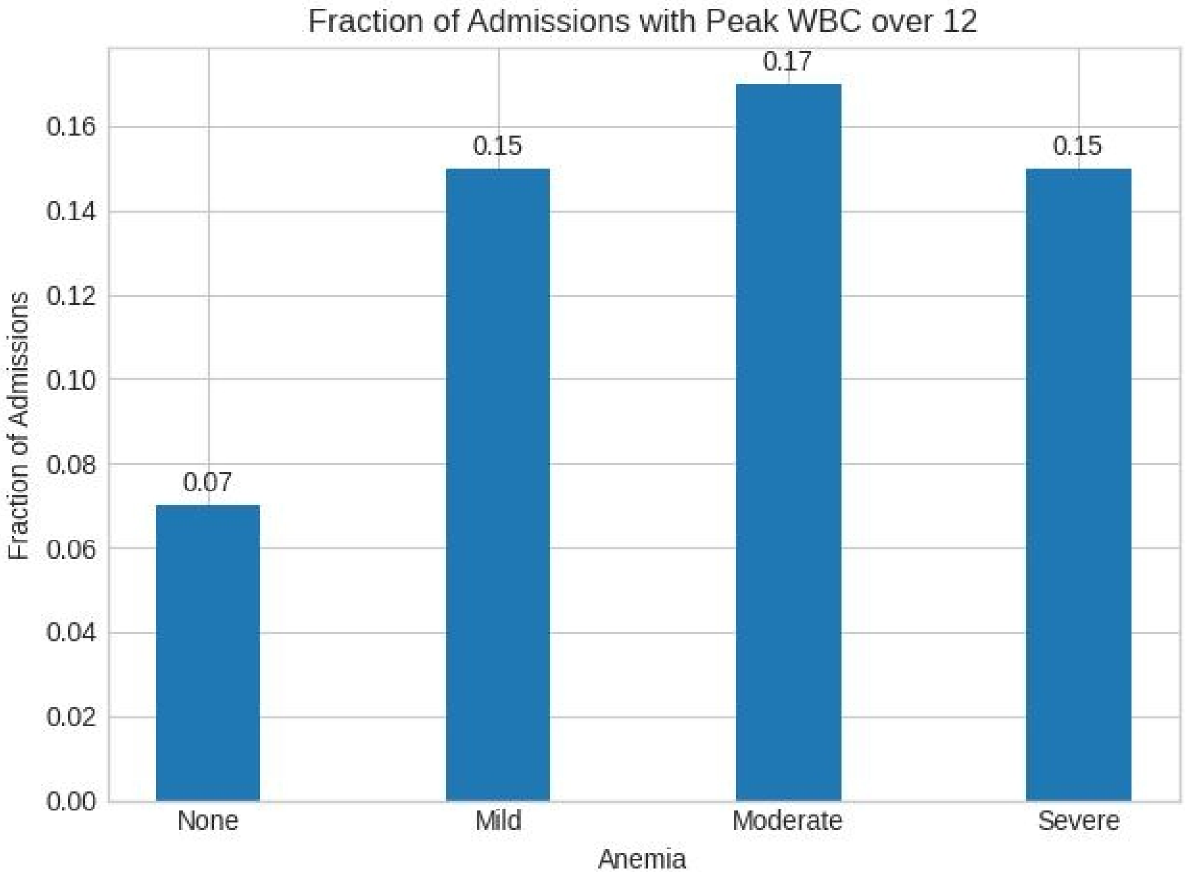


This is fraction of patients with each level of Hemoglobin Drop (not fraction of all admissions)

Figure 10 - Fraction Admissions with Anemia and Major Surgery

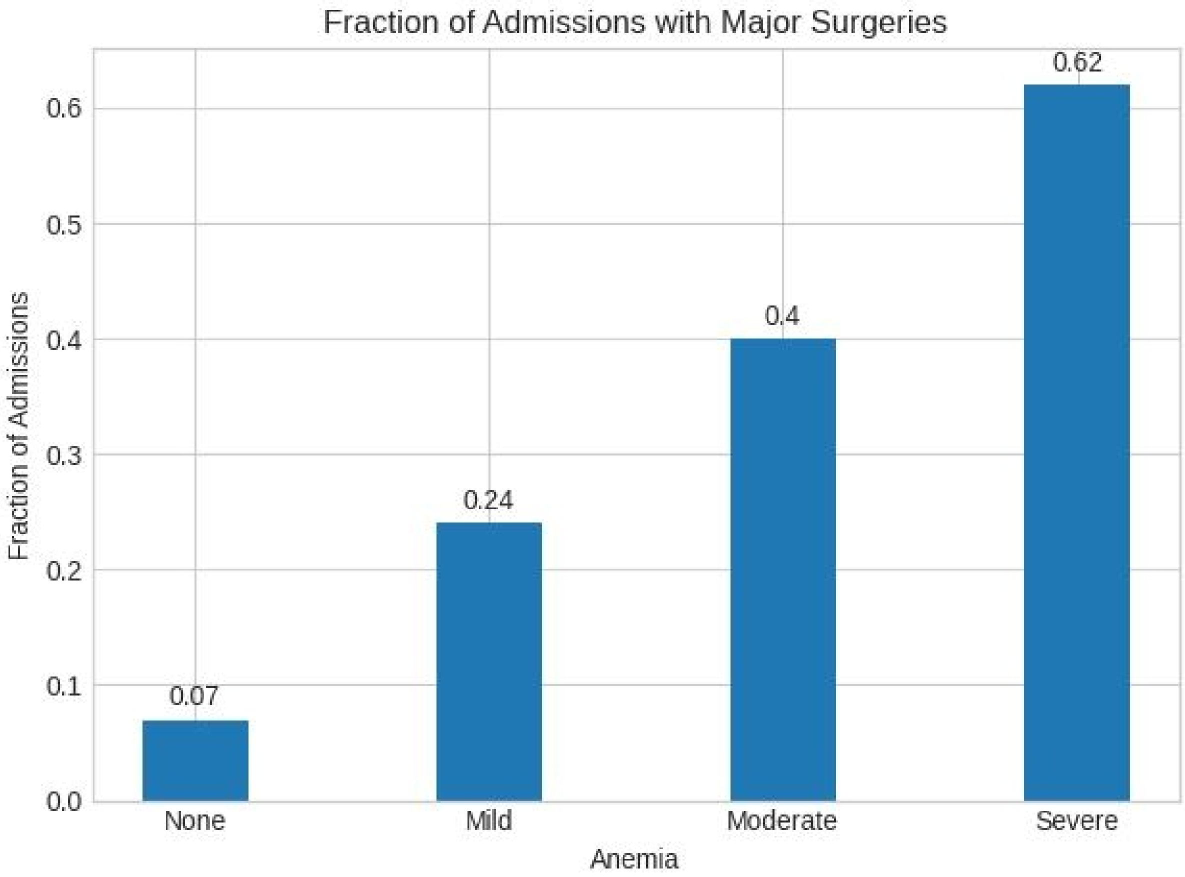


This is fraction of patients with each level of Hemoglobin Drop (not fraction of all admissions)

Figure 11

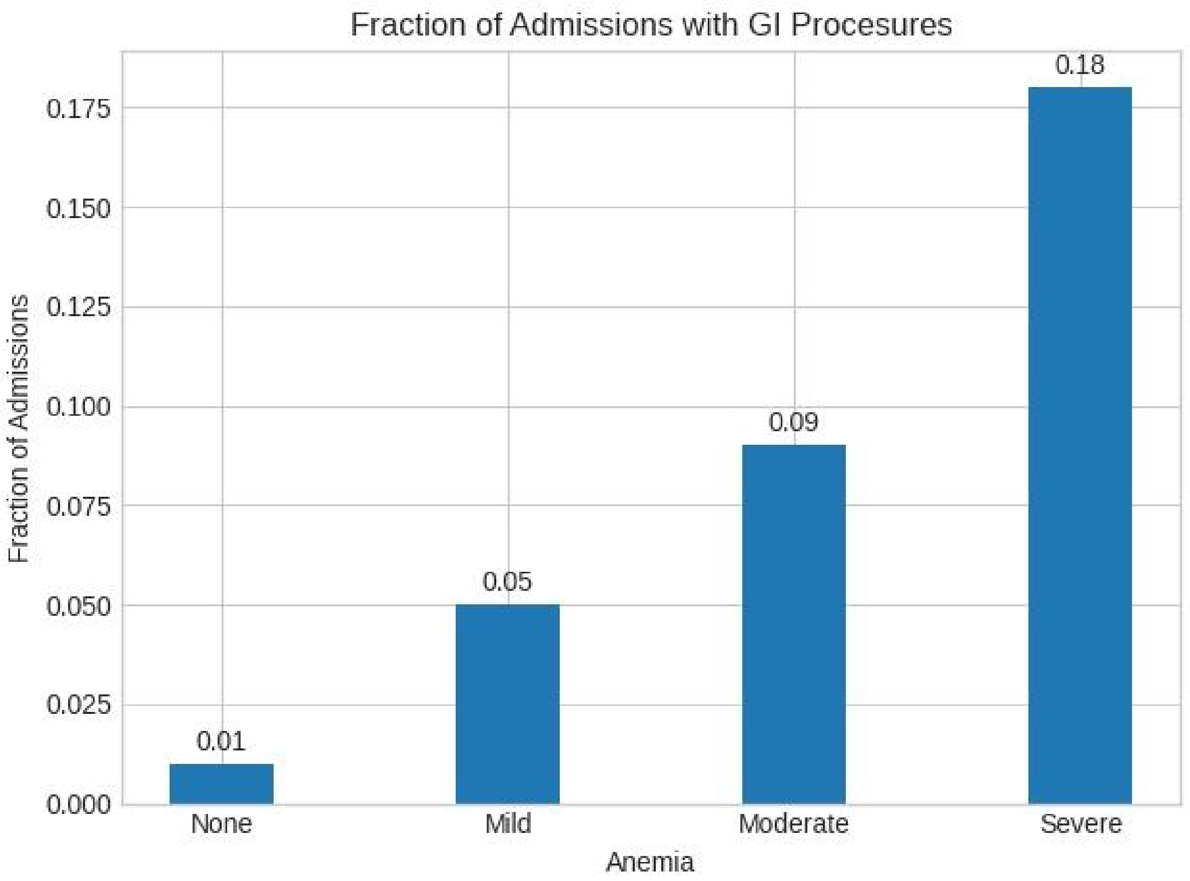


This is fraction of patients with each level of Hemoglobin Drop (not fraction of all admissions)

Figure 12

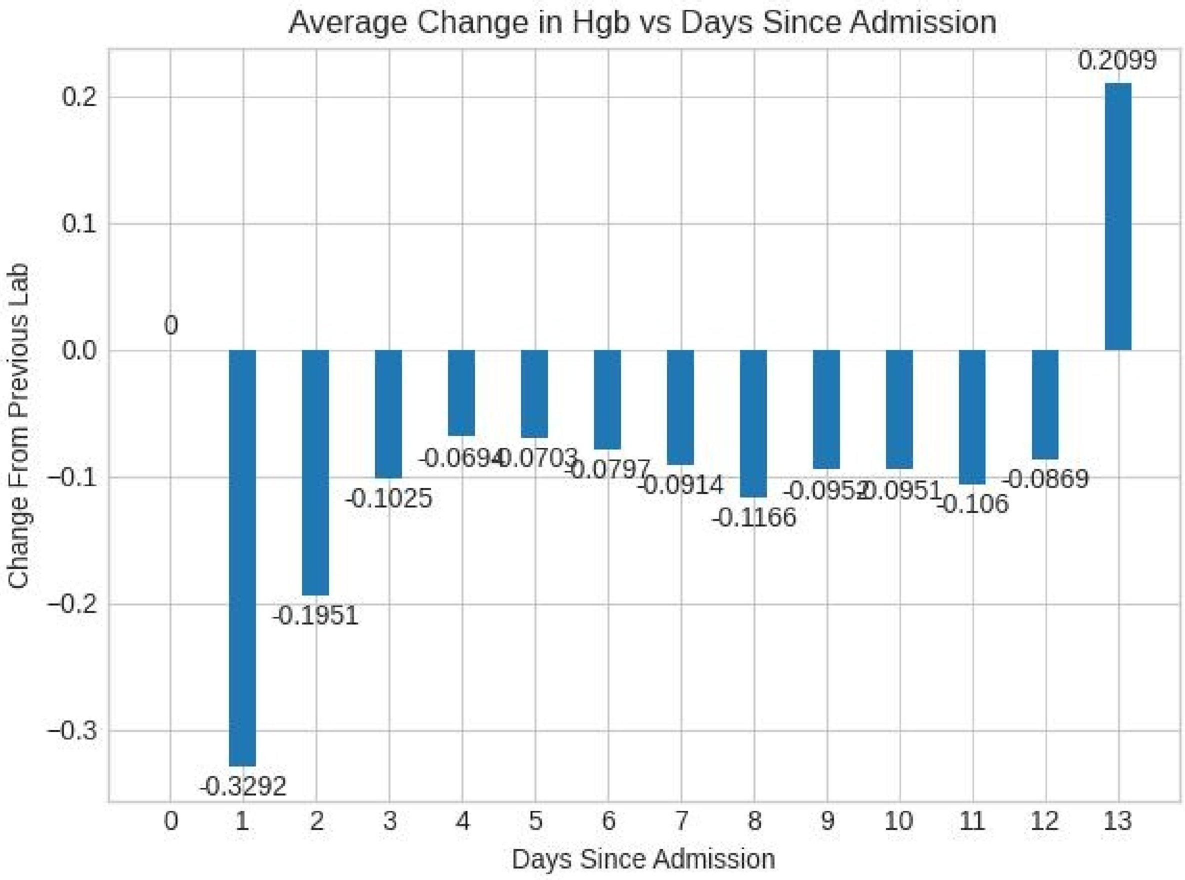
Average Change in Hgb vs Days Since Admission
Change From Previous Lab
Days Since Admission
0.2
0.1
0.0
−0.1
−0.2
−0.3
0
1
2
3
4
5
6
7
8
9
10
11
12
13
0
-0.3292
-0.1951
-0.1025
-0.0694
-0.0703
-0.0797
-0.0914
-0.1166
-0.0952
-0.0951
-0.106
-0.0869
0.2099

Figure 13

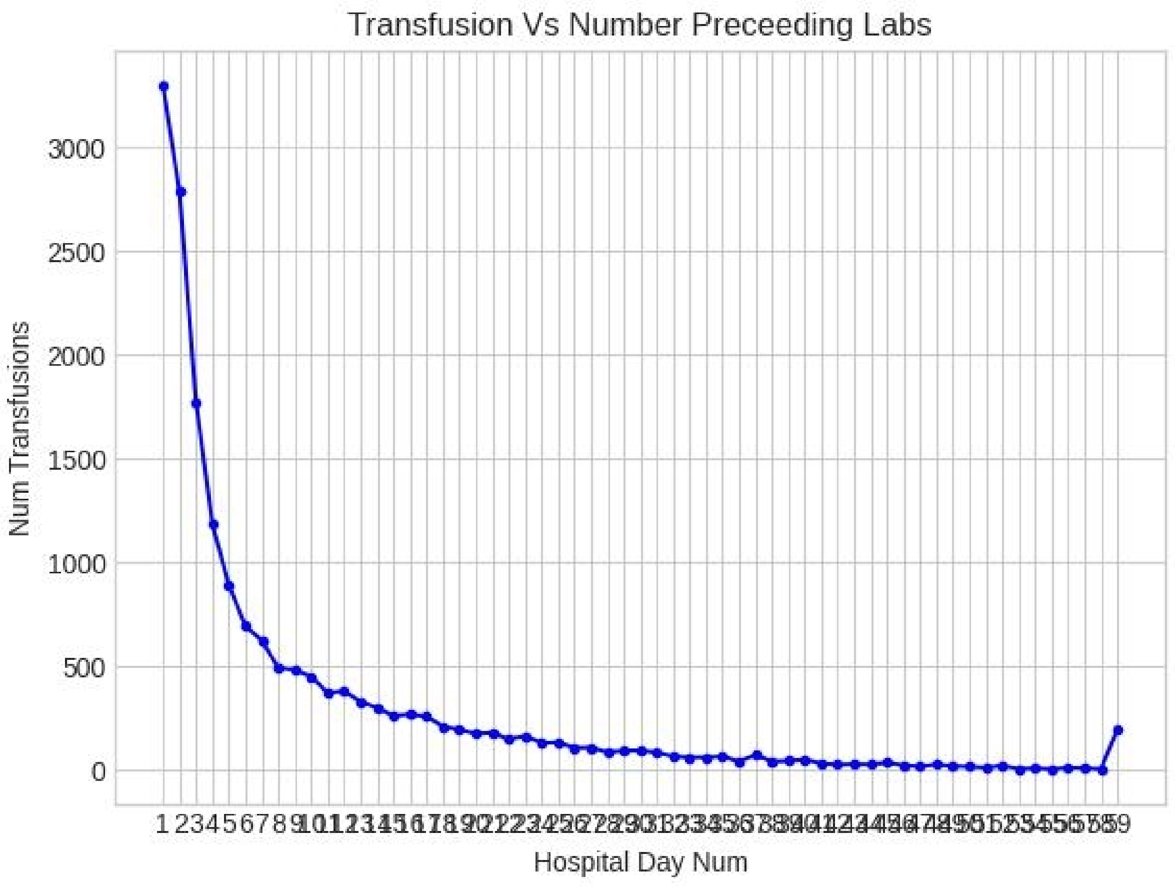

Transfusion Vs Number Preceeding Labs
Num Transfusions
Hospital Day Num
3000
2500
2000
1500
1000
500
0

Figure 14

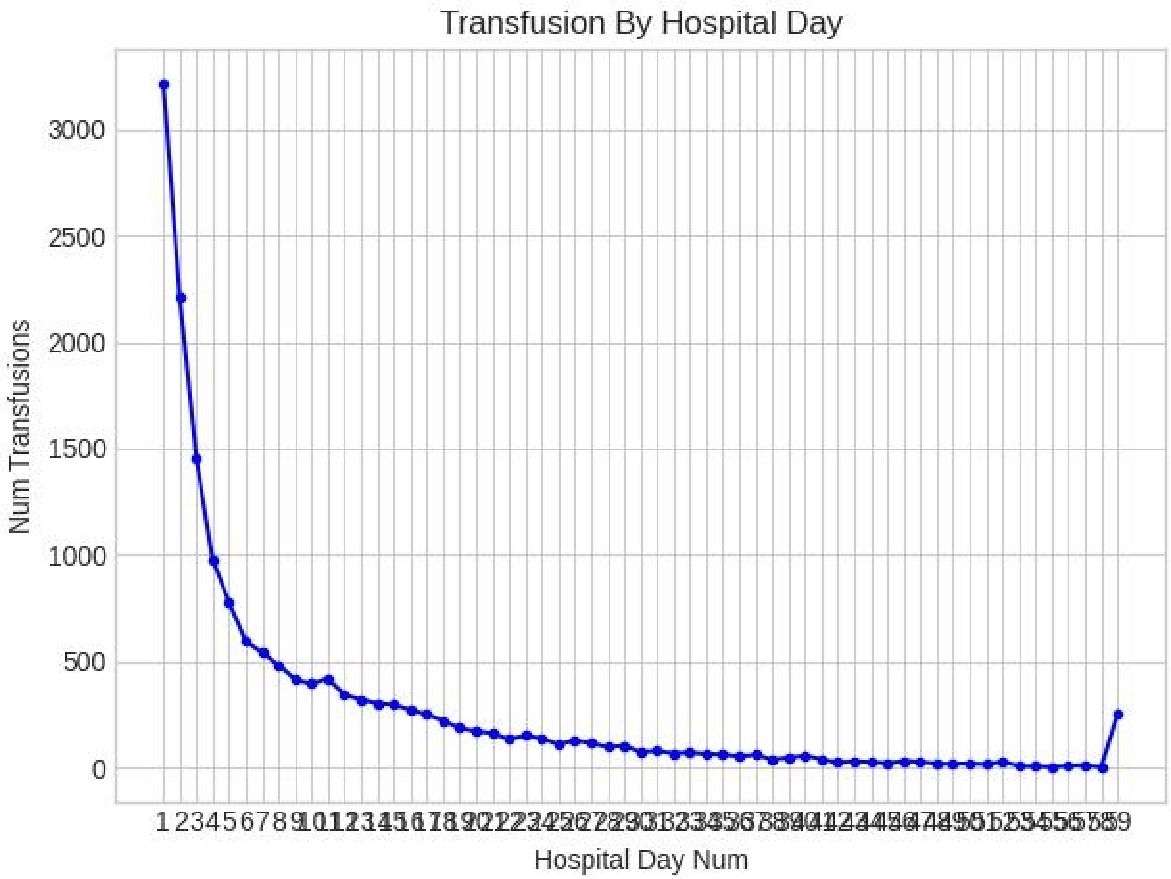
Transfusion By Hospital Day
Num Transfusions
3000
2500
2000
1500
1000
500
0
Hospital Day Num

Figure 15

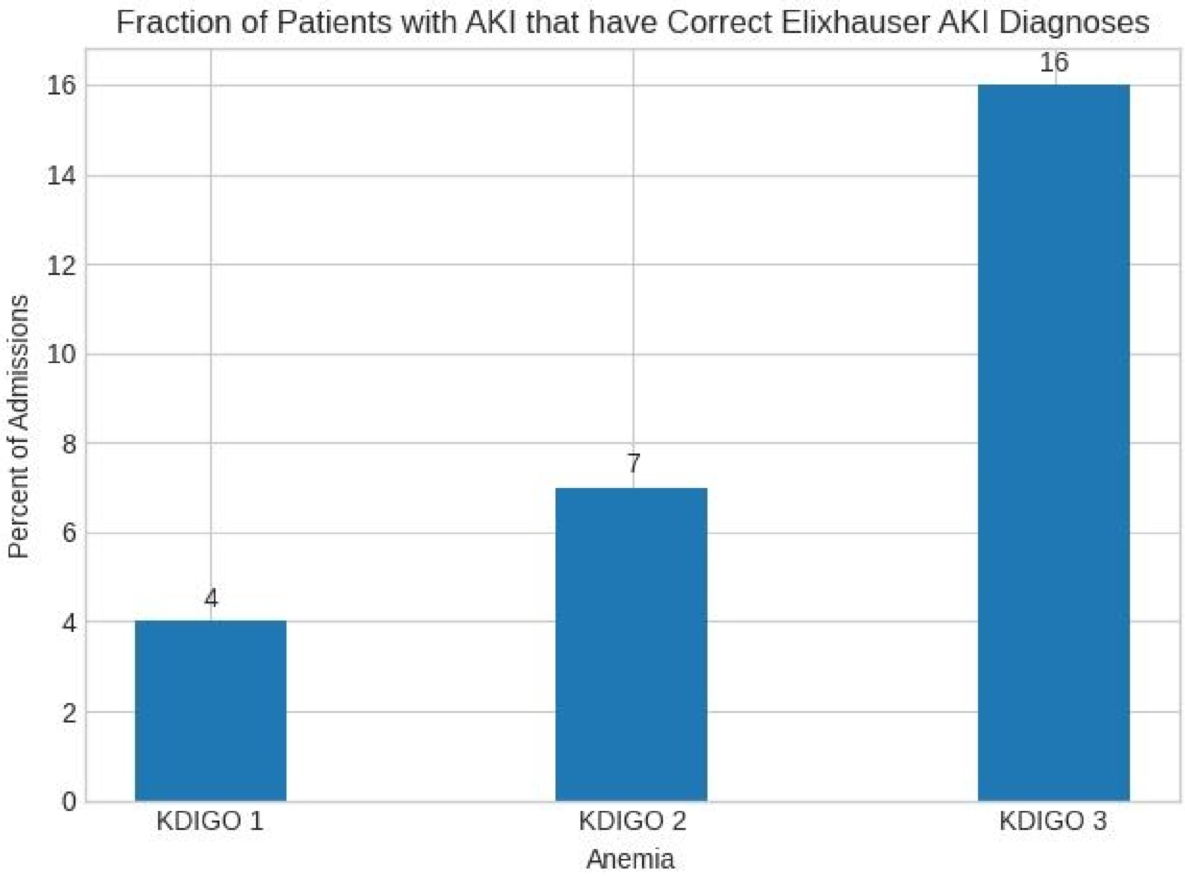

Fraction of Patients with AKI that have Correct Elixhauser AKI Diagnoses
Percent of Admissions
0
2
4
6
8
10
12
14
16
4
7
16
KDIGO 1
KDIGO 2
KDIGO 3
Anemia

Figure 16

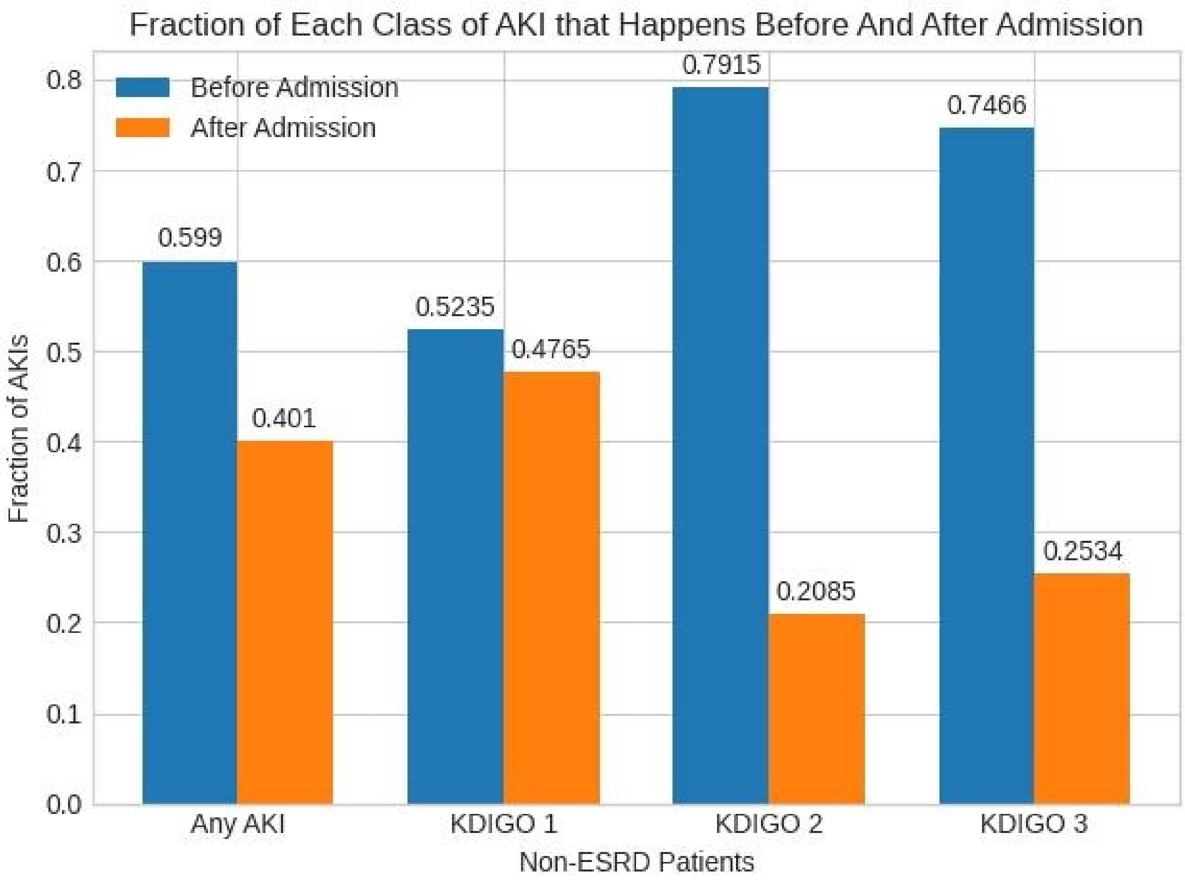
Fraction of Each Class of AKI that Happens Before And After Admission
Before Admission
After Admission
0.8
0.7
0.6
0.5
0.4
0.3
0.2
0.1
0.0
Fraction of AKIs
0.599
0.401
0.5235
0.4765
0.7915
0.2085
0.7466
0.2534
Any AKI
KDIGO 1
KDIGO 2
KDIGO 3
Non-ESRD Patients

Figure 17

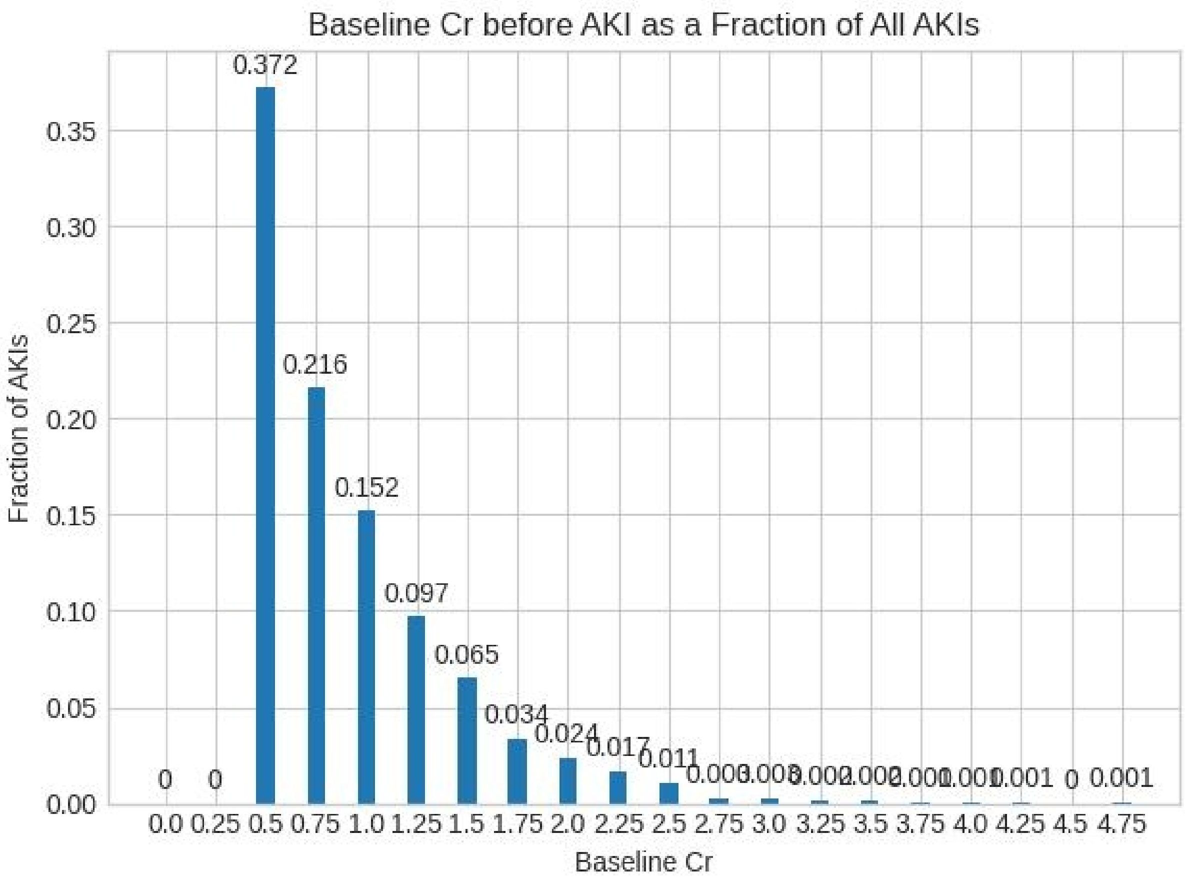
Baseline Cr before AKI as a Fraction of All AKIs
Fraction of AKIs
0.00
0.05
0.10
0.15
0.20
0.25
0.30
0.35
0.372
0.216
0.152
0.097
0.065
0.034
0.024
0.017
0.011
0
0
0.001
0
0.001
0.0 0.25 0.5 0.75 1.0 1.25 1.5 1.75 2.0 2.25 2.5 2.75 3.0 3.25 3.5 3.75 4.0 4.25 4.5 4.75
Baseline Cr

Figure 18

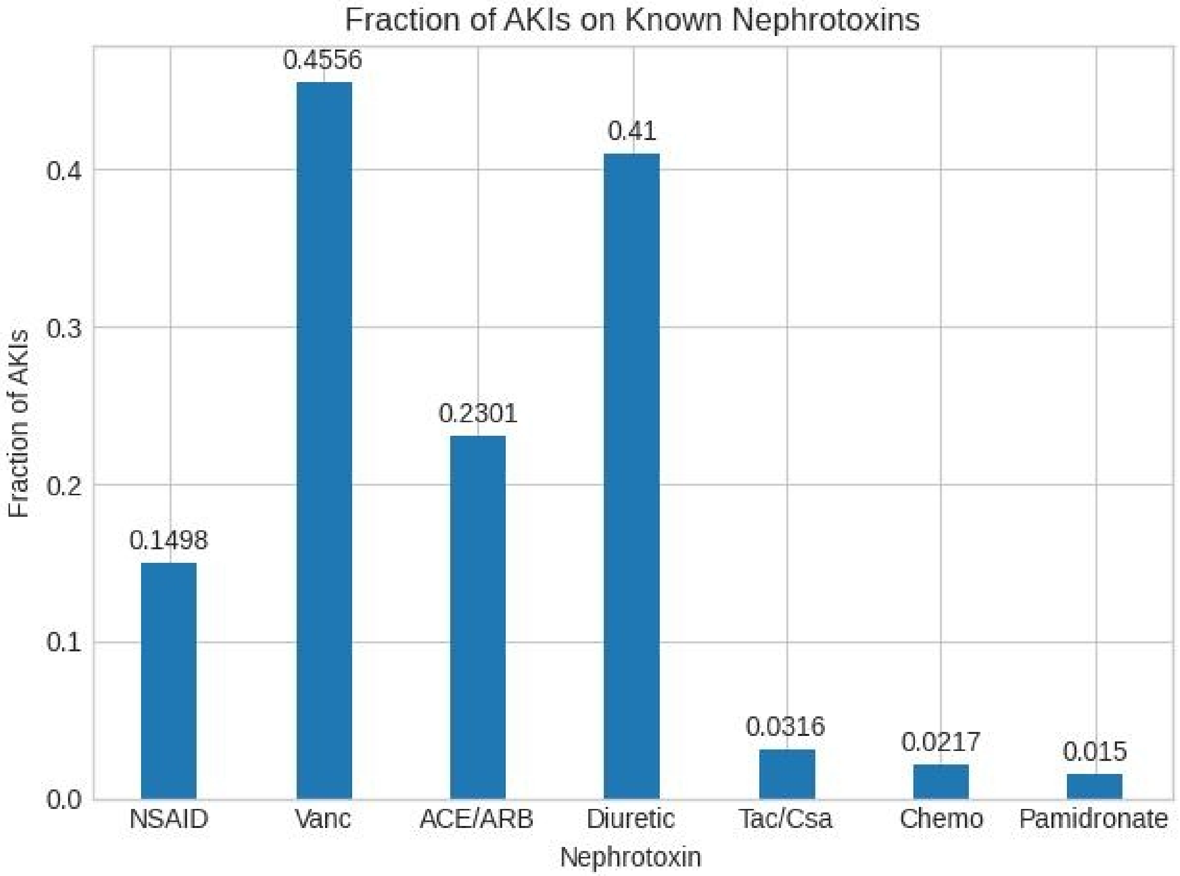
Fraction of AKIs on Known Nephrotoxins
Fraction of AKIs
Nephrotoxin
NSAID
Vanc
ACE/ARB
Diuretic
Tac/Csa
Chemo
Pamidronate
0.1498
0.4556
0.2301
0.41
0.0316
0.0217
0.015
0.0
0.1
0.2
0.3
0.4

Figure 19

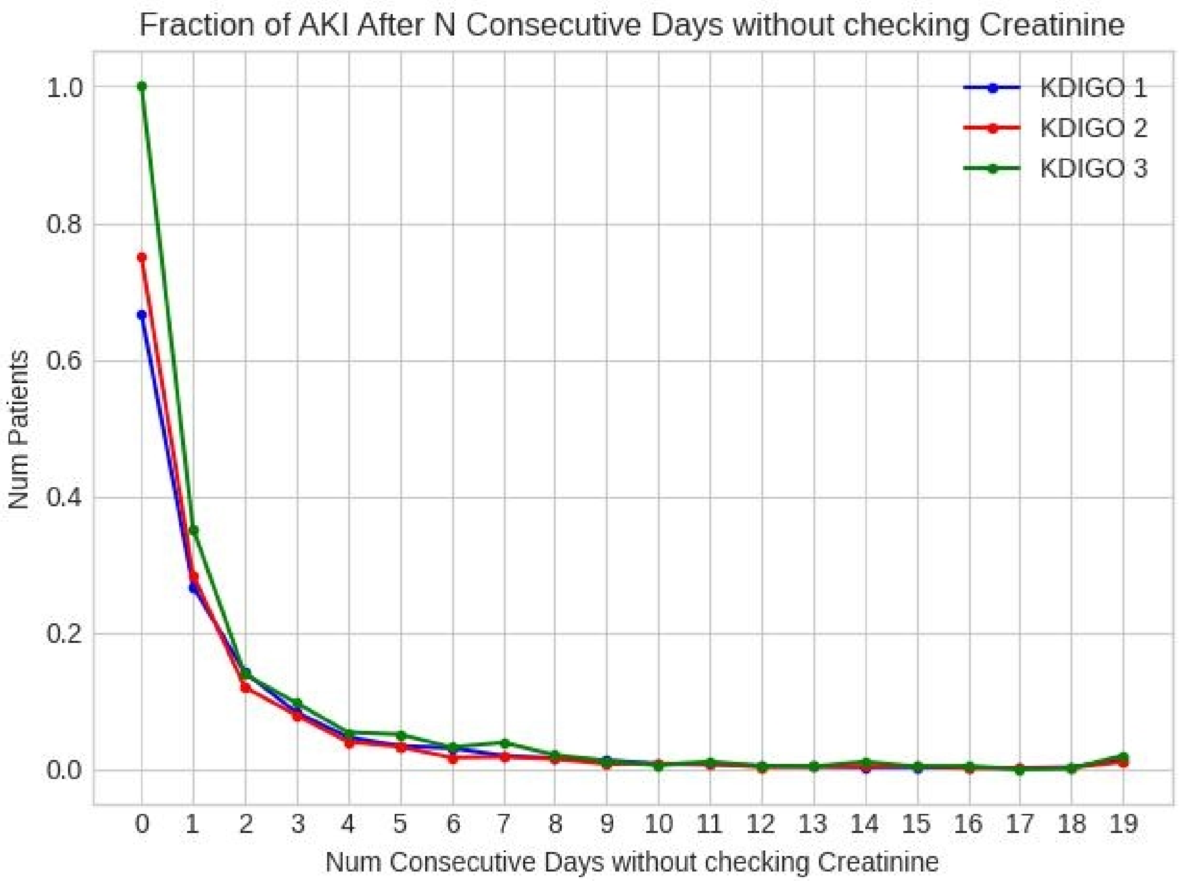

Fraction of AKI After N Consecutive Days without checking Creatinine
KDIGO 1
KDIGO 2
KDIGO 3
Num Patients
1.0
0.8
0.6
0.4
0.2
0.0
0 1 2 3 4 5 6 7 8 9 10 11 12 13 14 15 16 17 18 19
Num Consecutive Days without checking Creatinine

Figure 20 – Random Skipping

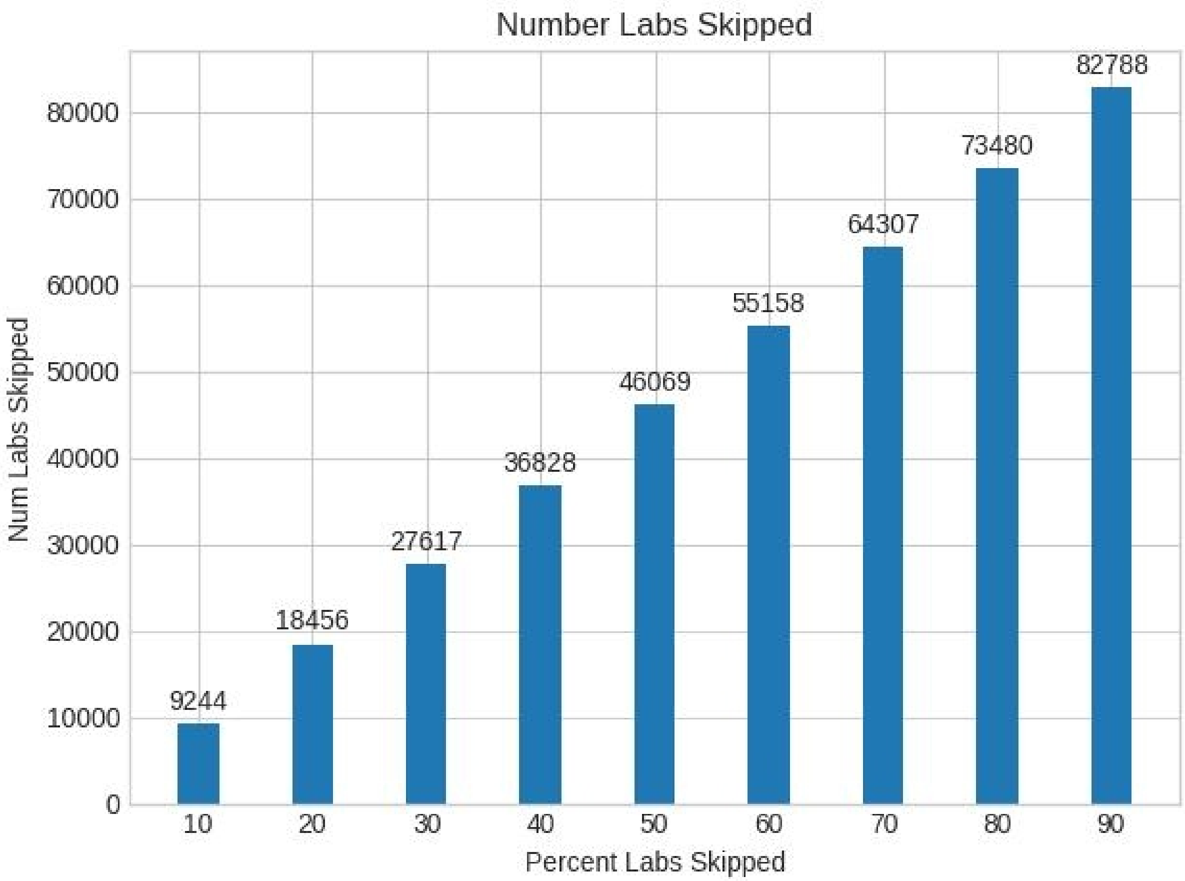

Figure 21 – Random Skip After 14 days

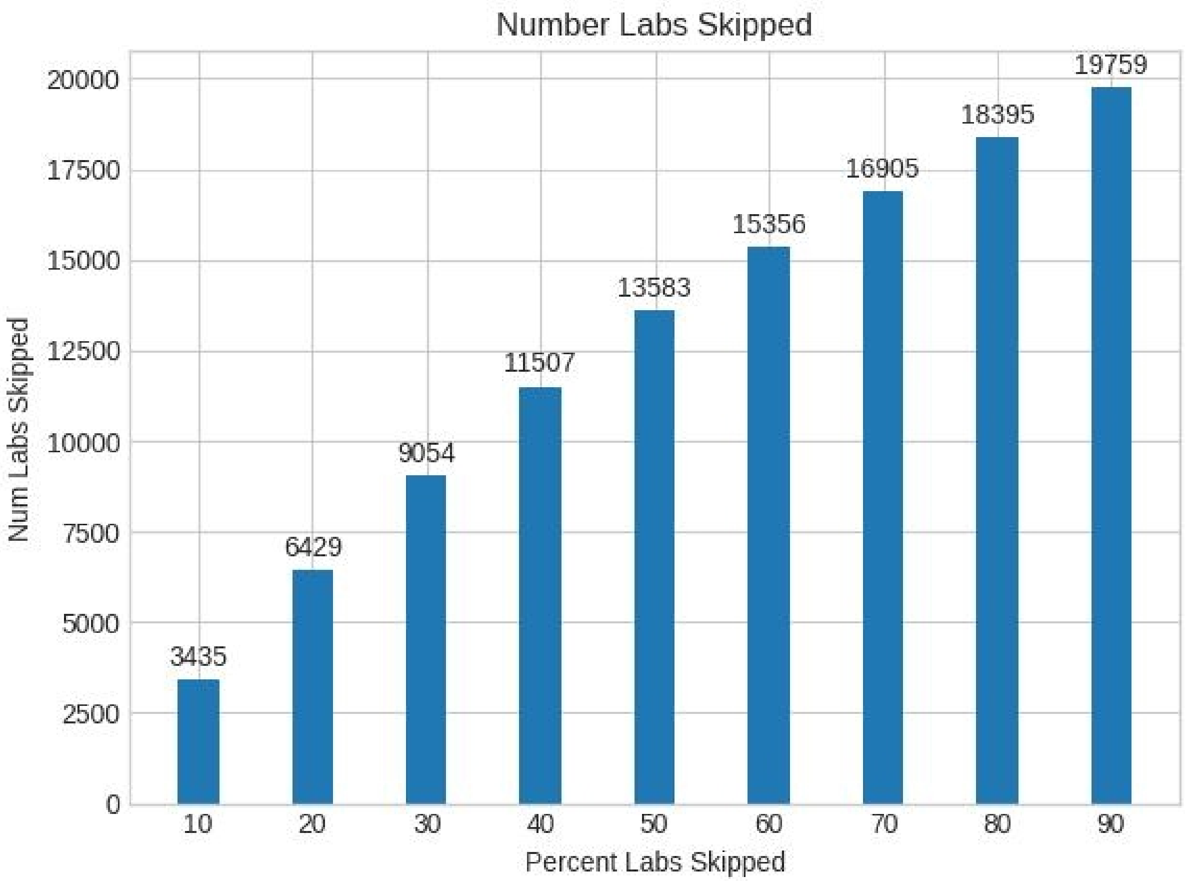

Figure 22

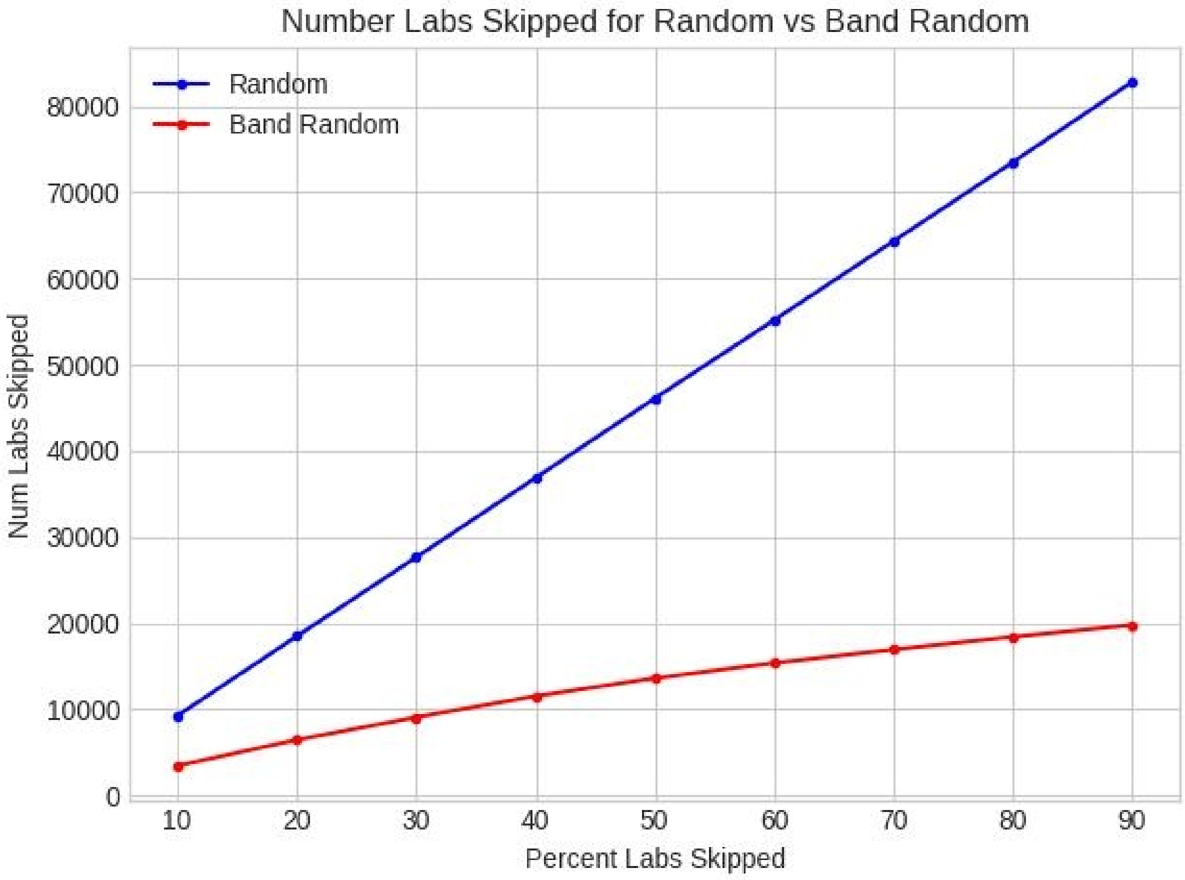
Number Labs Skipped for Random vs Band Random
Random
Band Random
Num Labs Skipped
Percent Labs Skipped
0
10000
20000
30000
40000
50000
60000
70000
80000
10
20
30
40
50
60
70
80
90

Figure 23

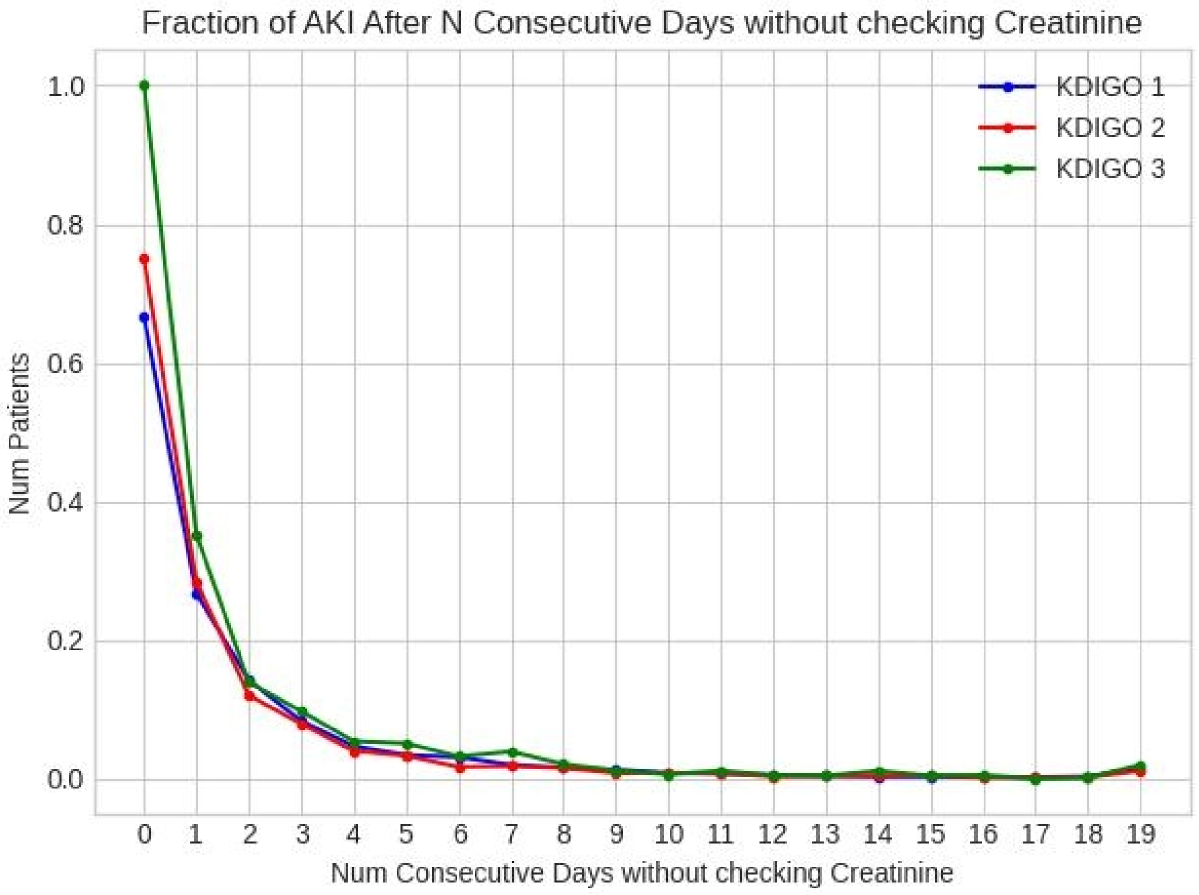

Fraction of AKI After N Consecutive Days without checking Creatinine
KDIGO 1
KDIGO 2
KDIGO 3
Num Patients
1.0
0.8
0.6
0.4
0.2
0.0
0 1 2 3 4 5 6 7 8 9 10 11 12 13 14 15 16 17 18 19
Num Consecutive Days without checking Creatinine

Figure 24

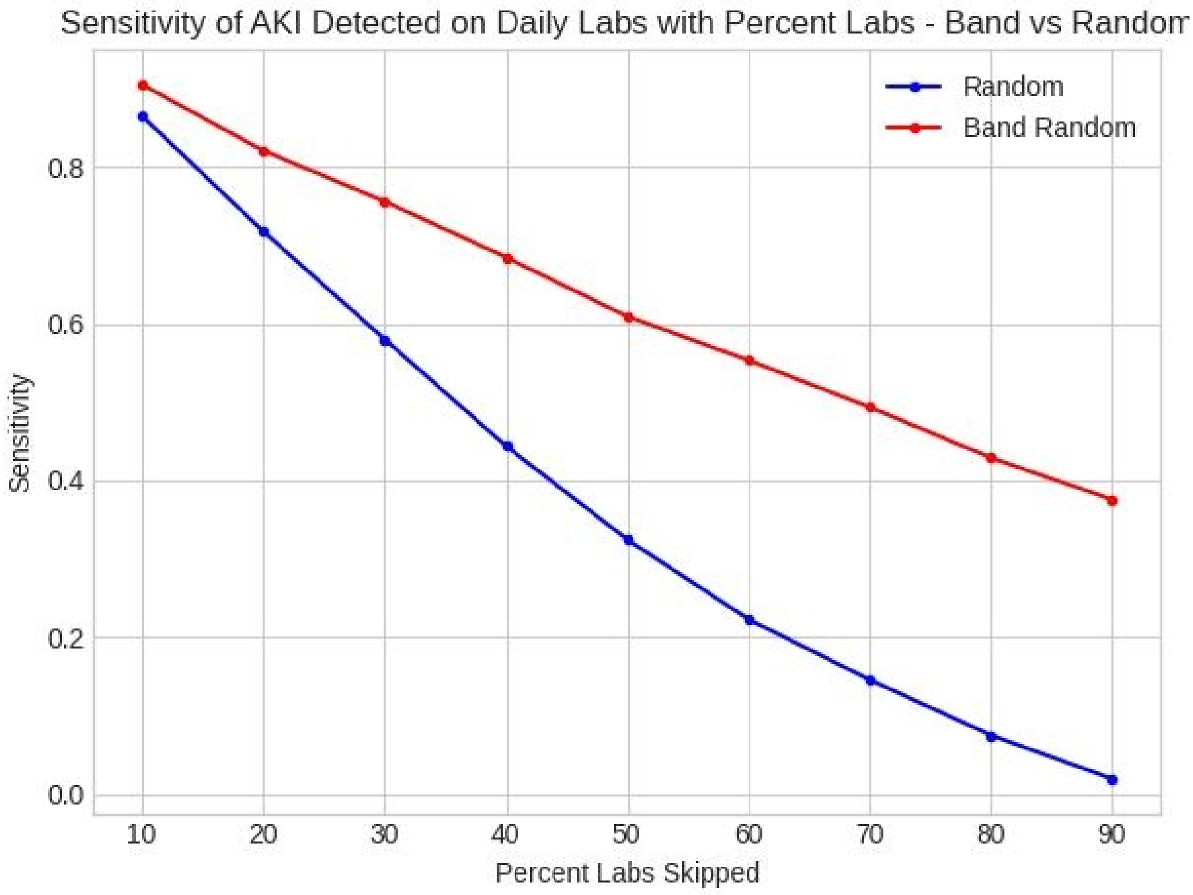

Sensitivity of AKI Detected on Daily Labs with Percent Labs - Band vs Random
Random
Band Random
Sensitivity
0.0
0.2
0.4
0.6
0.8
10
20
30
40
50
60
70
80
90
Percent Labs Skipped

Figure 25

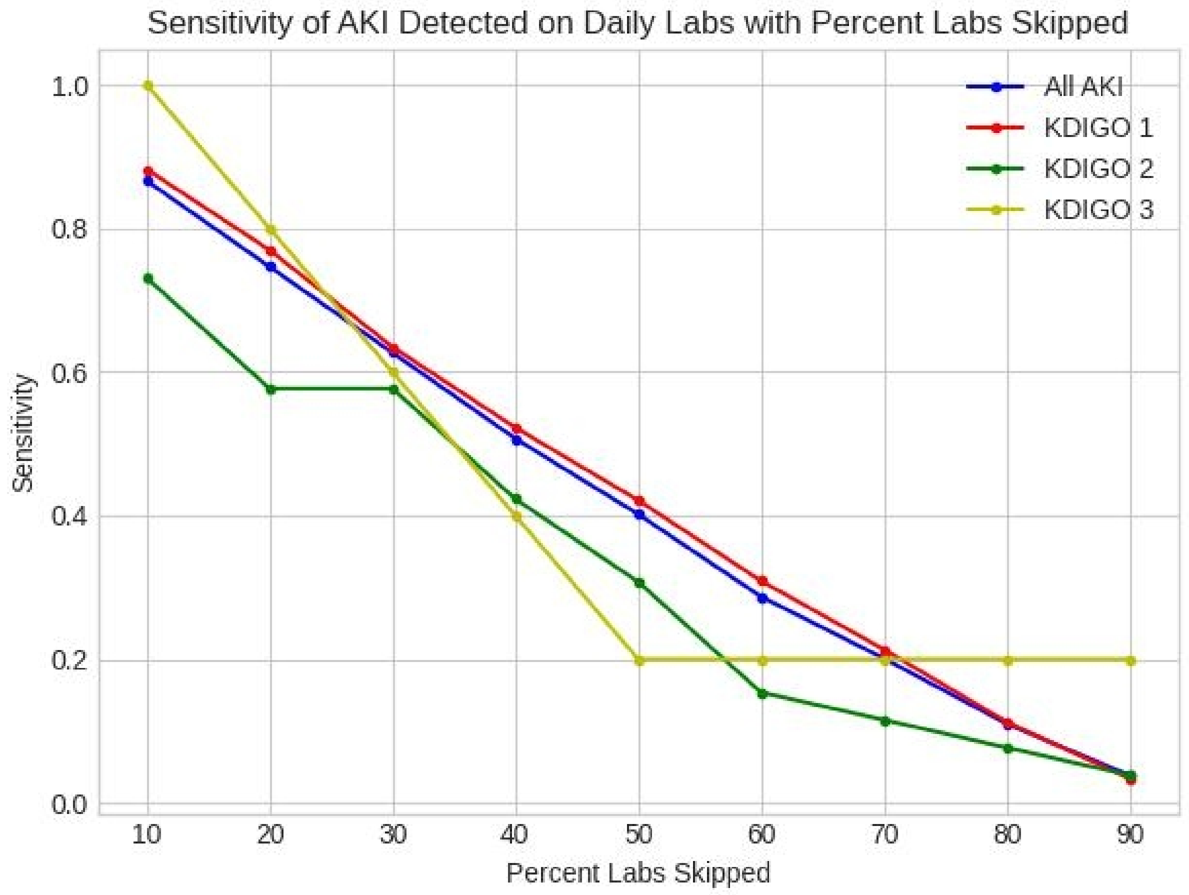
Sensitivity of AKI Detected on Daily Labs with Percent Labs Skipped
1.0
0.8
0.6
0.4
0.2
0.0
Sensitivity
10
20
30
40
50
60
70
80
90
Percent Labs Skipped
All AKI
KDIGO 1
KDIGO 2
KDIGO 3

Figure 26

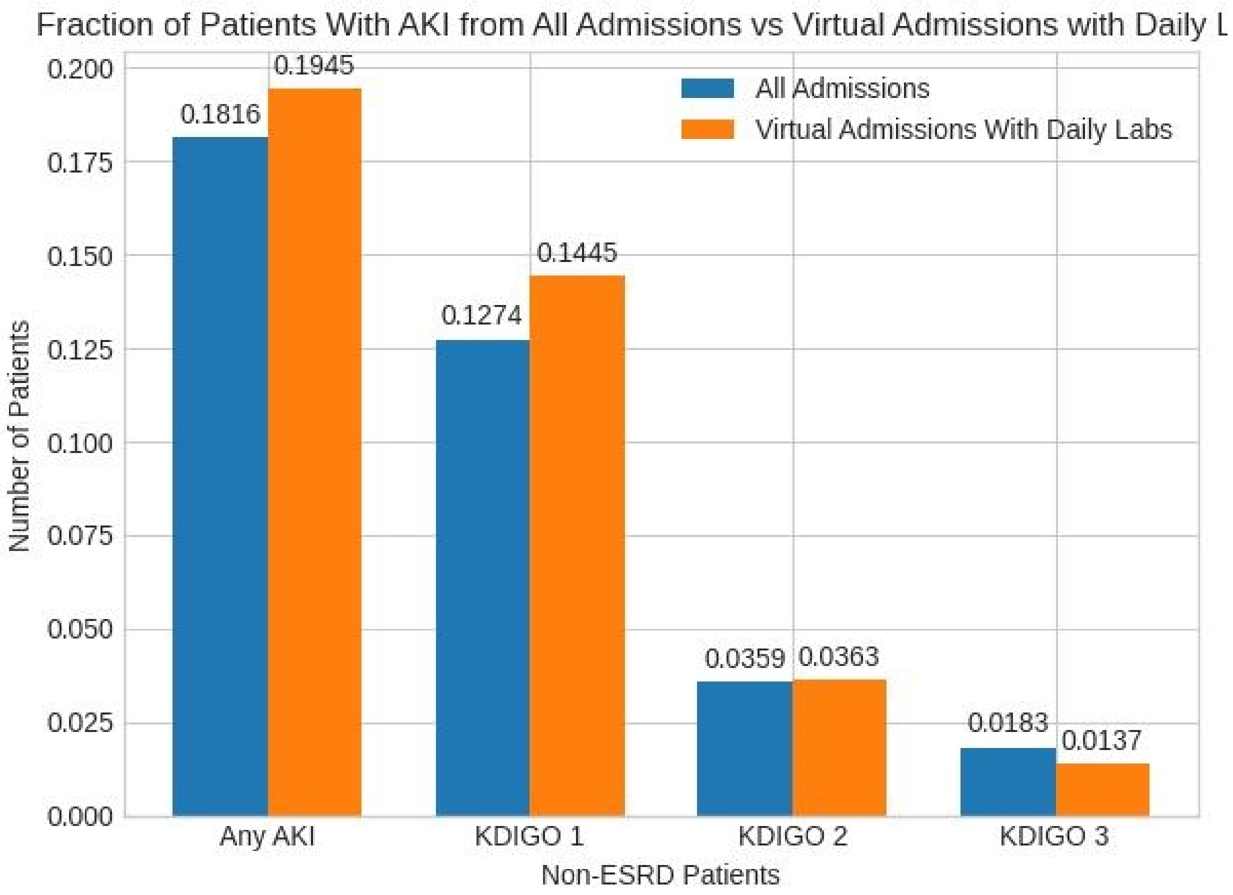
Fraction of Patients With AKI from All Admissions vs Virtual Admissions with Daily L
All Admissions
Virtual Admissions With Daily Labs
Number of Patients
0.200
0.175
0.150
0.125
0.100
0.075
0.050
0.025
0.000
0.1816
0.1945
0.1274
0.1445
0.0359
0.0363
0.0183
0.0137
Any AKI
KDIGO 1
KDIGO 2
KDIGO 3
Non-ESRD Patients

Figure 27

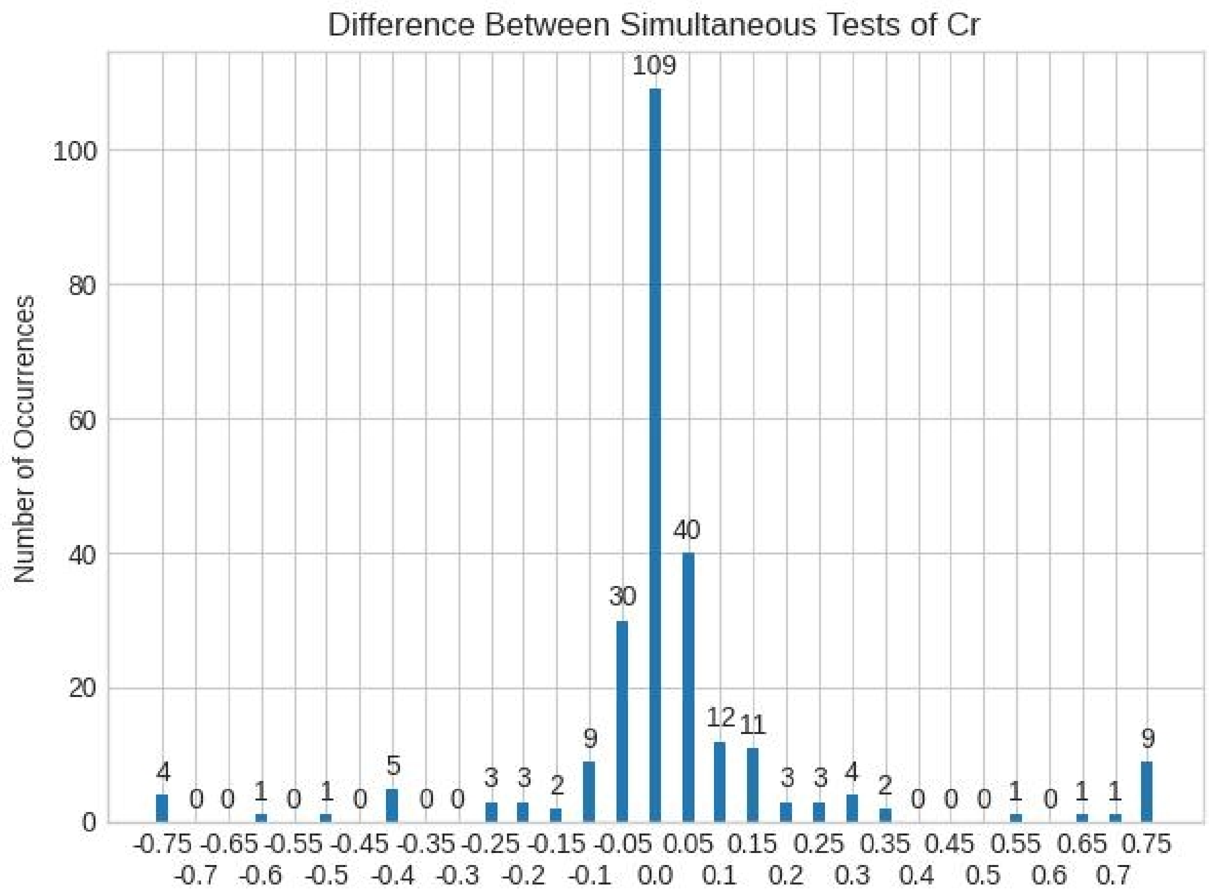

Difference Between Simultaneous Tests of Cr
Number of Occurrences
0
20
40
60
80
100
109
40
30
12
11
9
9
5
4
4
3
3
3
3
2
2
1
1
1
1
1
0
-0.75 -0.65 -0.55 -0.45 -0.35 -0.25 -0.15 -0.05 0.05 0.15 0.25 0.35 0.45 0.55 0.65 0.75
-0.7 -0.6 -0.5 -0.4 -0.3 -0.2 -0.1 0.0 0.1 0.2 0.3 0.4 0.5 0.6 0.7
